# Circulating extracellular vesicles in serum carry Trop2 marker for prostate cancer liquid biopsy and clinical care

**DOI:** 10.1101/2025.04.03.25325134

**Authors:** Prima Dewi Sinawang, Mehmet O. Ozen, Shiqin Liu, En-Chi Hsu, Demir Akin, Emily Ding, Rosalie Nolley, James D. Brooks, Tanya Stoyanova, Utkan Demirci

## Abstract

Extracellular vesicles (EVs) are lipid nano-to-micro-sized vesicles increasingly identified as valuable liquid biopsy tools for medical applications. However, the heterogeneity of cargo and the lack of convenient quantification methods to characterize EVs pose challenges in identifying vesicles with specific markers. In this study, we show the isolation, characterization, detection, and quantification of a cancer-specific marker, Trop2, on circulating extracellular vesicles in serum (EV-Trop2). This work combines the unique advantages of our user-friendly isolation method with serum diagnostics to identify high-risk prostate cancer cases and predict recurrence after prostate surgery. To our knowledge, this is the first demonstration to isolate and quantify EV-Trop2 from prostate cancer patient serum to study its analytical validity and potential clinical utility as an EV-based liquid biopsy. Initial study with patient serum samples from three clinical groups: high- risk prostate cancer (n = 22), low-risk prostate cancer (n = 23), and cancer-free groups (n = 21), demonstrates the potential of this approach in distinguishing prostate cancer aggressiveness. We observed significantly different levels of EV-Trop2 expression between the high-risk and low-risk patient groups (p = 0.0015), and between high-risk patient and cancer-free groups (p < 0.0001). Furthermore, employing machine learning algorithms, EV-Trop2 was shown to enhance classifier metrics across the three sample groups, aiding both in risk stratification and predicting recurrence post-prostatectomy. The availability of such tool could have a broad impact across multiple cancers by enabling minimally invasive liquid biopsy sampling.

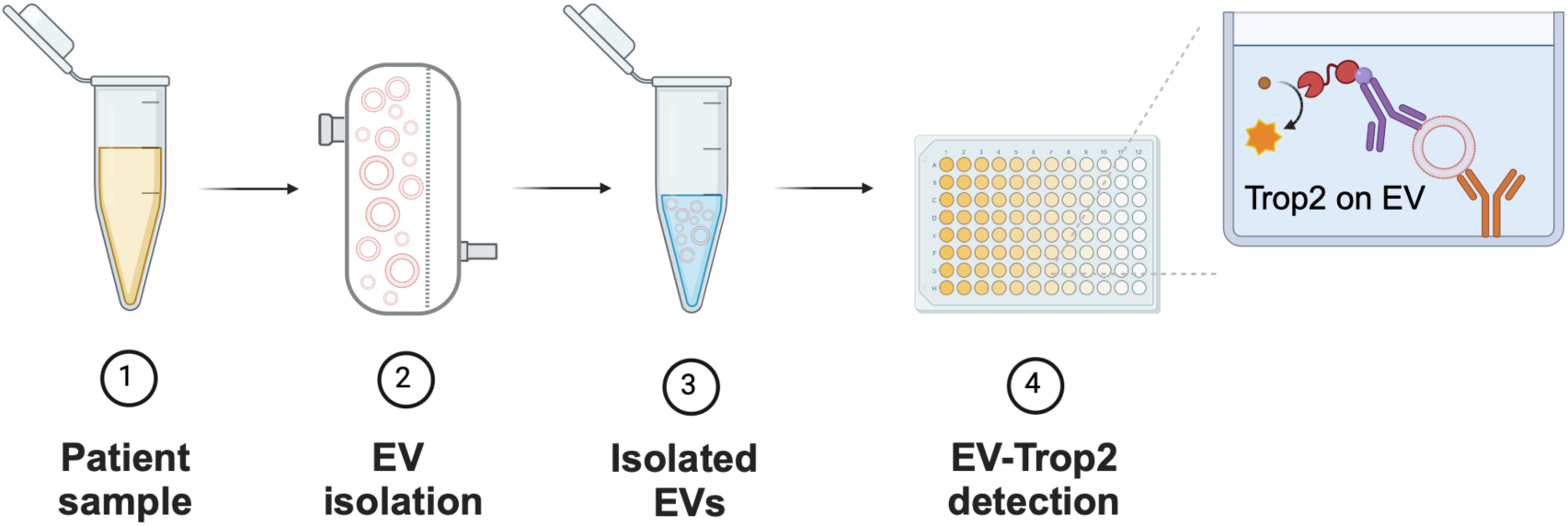

## INTRODUCTION

Tissue biopsies and imaging play a pivotal role in cancer diagnosis, but are associated with invasive procedures, patient discomfort, and radiation exposure.^1,2^ These drawbacks underscore the appeal of liquid biopsy-based analyses, which offer minimal invasiveness, real-time monitoring capabilities, and enhanced genomic and proteomic resolution, driven by advancements in recent years.^3,4^ Such tools present a promising complementary minimally invasive approach that could be used before ordering tissue biopsy.

Extracellular vesicles and particles (EVPs) and their characterizations in clinical samples hold immense potential for driving advancements with broad applicability in various fields, including diagnostics.^5^ Exosomes, a well-known subclass of EVPs, are small lipid bilayer structures measuring smaller than 200 nm in diameter, originating from the inward budding of endosomal membranes.^6,7^ However, current standard isolation techniques have limitations in definitively enriching and characterizing EVPs based on their biogenesis mechanisms.^7,8^ To ensure consistency in terminology throughout this paper, we adopt the generic and operational term “EVs”, which has been used over the last few years to denote EV populations in general.^7^ EVs offer unique opportunities for liquid biopsy applications, particularly because they can be found in clinical samples (*e.g.,* whole blood) alongside other components^9^, including circulating tumor cells (CTCs),^10^ circulating tumor DNA (ctDNA) or cell-free DNA (cfDNA),^11^ and other circulating RNAs^12^ and proteins.^13^ CTCs, while informative, suffer from being rare,^14^ and cfDNA’s short half-life limits its detectability.^15^ In contrast, EVs can be abundant, numbering in the billions to trillions (10^10^–10^12^) per milliliter of plasma.^16^ These EVs are secreted by a wide range of cell types, including cancer cells.^17^ They are present in various bodily fluids, including blood, saliva, urine, and lavage.^18^ They serve as intercellular transport vehicles, ferrying a cargo of molecules within or on the surface, such as RNA,^19,20^ DNA,^21,22^ proteins,^23,24^ lipids,^25,26^ and metabolites^27,28^ from donor cells to recipient cells and tissues. As a result, EVs bear specific signatures and biomarkers that can reflect the disease state and progression of the originating cancer cells, making them promising targets for early diagnosis and prognosis assessment.^29^ However, identifying vesicles with specific markers remains difficult due to the heterogeneous nature of EV cargo.^30,31^

In this study, we demonstrate the isolation, characterization, and application of EVs for prostate cancer detection and clinical care, as prostate cancer is the most commonly diagnosed cancer in males in 118 of 185 countries,^32^ and the second leading cause of cancer-associated deaths in men in the United States.^33,34^ Our study focuses on serum-derived EVs carrying Trop2 on their surfaces. Trop2, an oncogenic protein (encoded by *TACSTD2* gene), has emerged as a promising marker due to its differential expression across various epithelial cancers.^35^ In the context of prostate cancer, Trop2 is elevated compared to normal tissue, and only 10% of prostate adenocarcinomas are Trop2-negative.^36–38^ Trop2 is a transmembrane protein implicated in various cellular processes (*e.g.,* apoptosis inhibition, cell proliferation, cell migration, and metastasis).^39^ High Trop2 levels are correlated with worse overall survival and a higher risk of recurrence in prostate cancer.^36,38^ While normal cells can indeed express Trop2 on their membranes,^40^ the overexpression of Trop2 protein on cancer cell membranes is also reflected on the membranes of EVs released by cells,^41^ referred to as EVs carrying Trop2 on the surface (EV-Trop2) (**Figure 1A**). This mechanism renders EVs as compelling and viable carriers of Trop2, presenting a promising liquid biopsy diagnostic modality as a signature of cancer in circulation. This concept was implemented in our study, where we enriched EVs from serum samples using our isolation platform (ExoTIC),^42^ providing a cleaner sample for more specific and accurate EV-Trop2 ELISA results.

**Figure 1.**
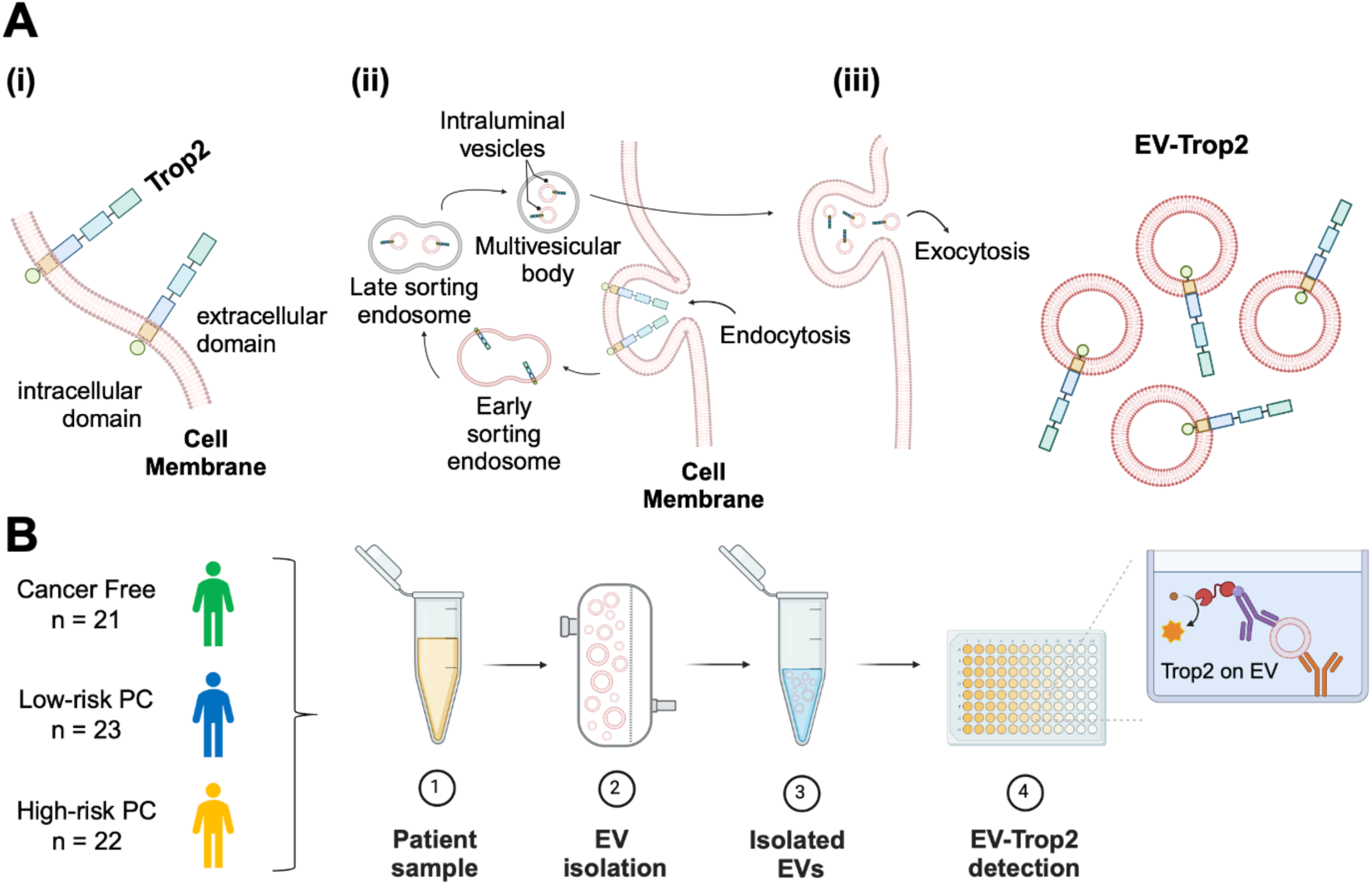
Development of a tool and workflow to isolate, characterize, and quantify Trop2 on the surface of serum-derived EVs from clinical prostate cancer samples. **(A)** Schematic representation of Trop2 distribution during extracellular vesicle (EV) biogenesis. (i) Trop2, a transmembrane glycoprotein overexpressed on the cancer cell surface,^36,38,43^ (ii) becomes enclosed within intraluminal vesicles during multivesicular body formation.^29^ (iii) The process culminates in exocytosis, releasing EVs^29^ decorated with overexpressed Trop2 (EV-Trop2). **(B)** Schematic representation of the workflow for EV-Trop2 isolation and detection. Clinical serum samples undergo isolation and enrichment of EVs using our well-established and well-published tool, ExoTIC.^42,44–47^ The Enzyme-Linked Immunosorbent Assay (ELISA) is employed for the detection of EV-Trop2, providing a user-friendly and scalable approach. The combination of both can potentially be leveraged for high-throughput diagnostics in the future. Figures were created with BioRender.com.

In this paper, we present the isolation and characterization of EVs accompanied by the quantification of Trop2 on the surface of EVs (EV-Trop2) in serum samples from three different clinical groups: high-risk prostate cancer (n = 22), low-risk prostate cancer (n = 23), and cancer-free groups (n = 21), demonstrating its analytical validity and potential clinical utility in prostate cancer (**Figure 1B**). To our knowledge, this is the first demonstration to isolate and quantify EV-Trop2 from prostate cancer patient serum. Our findings show that serum EV-Trop2 levels are different among the three different groups. Moreover, incorporating the EV-Trop2 data alongside the current standard, prostate-specific antigen (PSA) data, in multivariate analysis improves the cancer risk classification and recurrence prediction. Therefore, this tool could potentially be used as a complementary liquid biopsy tool to PSA testing to identify patients at risk for aggressive prostate cancer and predict recurrence.

## RESULTS AND DISCUSSION

In this study, we measured serum EV-Trop2 levels using ELISA. Using serum samples directly in the assay without pre-processing steps to eliminate background protein contaminants might result in a high background signal, potentially leading to indistinguishable results between cancer and cancer-free samples. To mitigate the effect of background protein contaminants, we focused on isolating and enriching EVs, which are recognized as intercellular transport vehicles, and have been demonstrated *in vitro* to transport Trop2 through extracellular secretion.^41^ The initial optimization of our ELISA assay involved using EVs from DU145, a prostate cancer cell line recognized for its elevated cellular Trop2 levels.^38^ Subsequently, we evaluated EV-Trop2 levels in clinical serum samples of prostate cancer patients.

We evaluated EV-Trop2 as a potential blood-based biomarker for clinically significant prostate cancer by isolating serum-derived EVs using our developed filtration-based tool, ExoTIC,^42^ followed by ELISA. Our EV-Trop2 isolation and detection workflow is outlined in **Figure 1B**. We conducted a comparative analysis of EV-Trop2 levels in three clinical sample groups (**Figure 2**): cancer-free individuals (biopsy-confirmed absence of prostate cancer), patients with low-risk prostate cancer (no biochemical recurrence for at least 5 years after prostate surgery), and those with high-risk prostate cancer (with biochemical recurrence after prostate surgery). The stratification of the patients was based on the National Comprehensive Cancer Network (NCCN) guidelines.^48^ Patient information including pre-operative PSA levels and %Gleason score (Grade 4 or 5) in the pathological specimen from the radical prostatectomy are summarized in **Supplementary Table S1**. We have broken down samples by the percentage of Gleason patterns 4 or 5 because data from our institution demonstrates a strong correlation with adverse outcomes (recurrence after surgery).^49^

**Figure 2.**
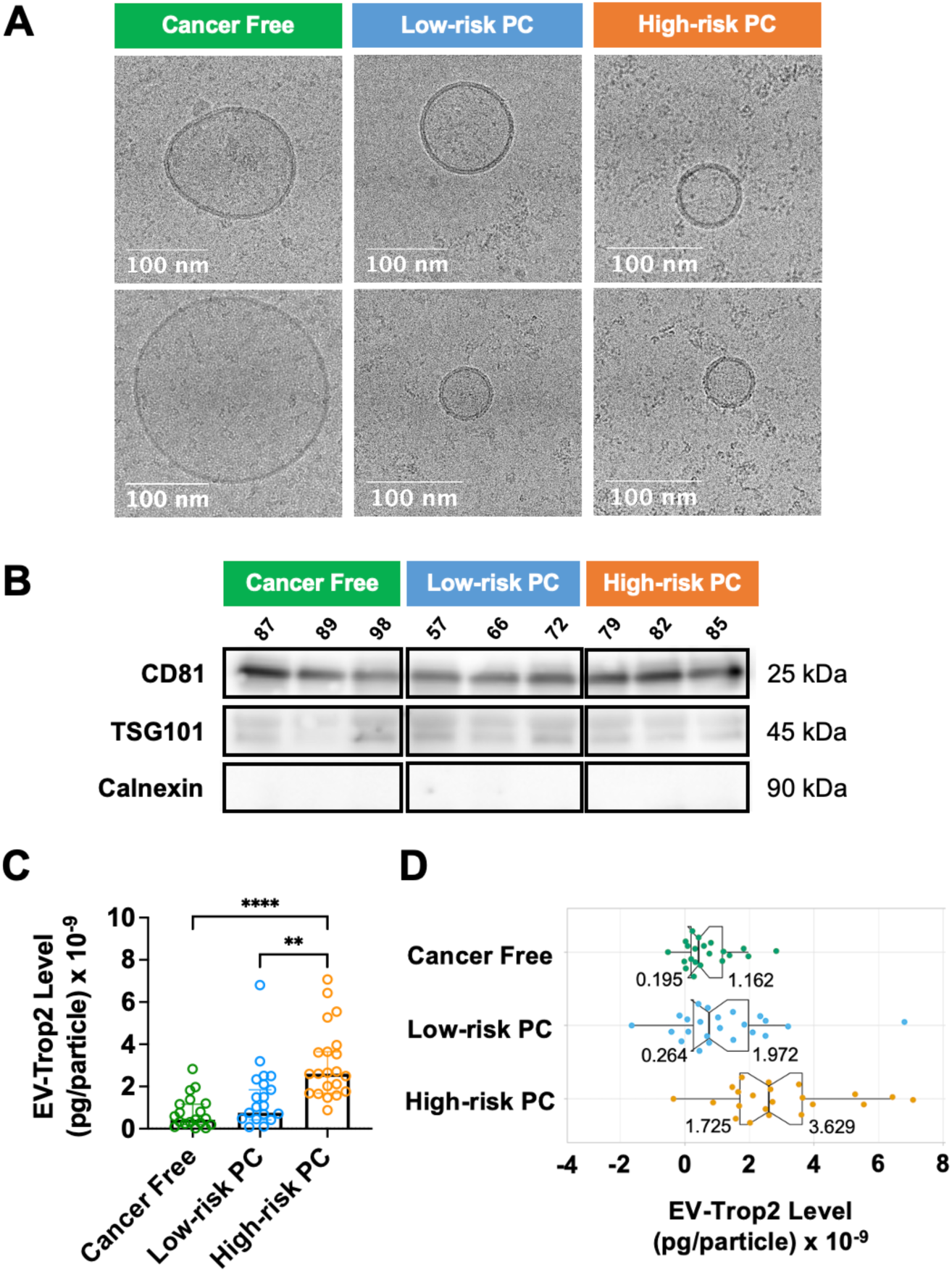
Profiling EVs from clinical serum samples. **(A)** Representative cryo-EM images of EVs from different sample groups (Scale bar: 100 nm), which exhibit typical EV-like structures. **(B)** Western Blot analysis of EVs using positive protein markers (CD81, TSG101) and negative marker (Calnexin). **(C)** Normalized EV-Trop2 levels across distinct clinical sample groups (cancer free, low-risk PC, high-risk PC) relative to corresponding EV numbers (measured by NTA). Statistical analysis: One-way ANOVA with Kruskal-Wallis’s multiple comparison test; ** p ≤ 0.01, **** p ≤ 0.0001. **(D)** Box plots illustrating the distribution of EV-Trop2 levels in each sample group. Lines are median values (Q2). 25% and 75% percentiles are indicated accordingly. Error bars are standard deviations.

### Characterization of serum-derived EVs from clinical samples

Through cryogenic transmission electron microscopy (cryo-EM), we observed that the isolated EVs from serum exhibited typical EV-like structures. These structures appeared intact, circular, and were characterized by a lipid bilayer membrane configuration across all samples (**Figure 2A**). The size distributions of the isolated EVs, measured using Nanoparticle Tracking Analysis (NTA), are presented in **Supplementary Figure S1**, where the mean and median sizes across the different clinical sample groups were observed to be smaller than 200 nm. According to the NTA data, there was no statistically significant difference in EV size between cancer- free (n = 12) and low-risk PC (n = 13) samples. However, high-risk PC samples (n = 13) showed significantly smaller EV sizes compared to the other two groups. This finding aligns with previous reports showing that more aggressive forms of cancer tend to be associated with smaller EV sizes.^50^ While this observation is promising, it requires further validation in a larger cohort. Moreover, while our isolated EVs might include lipoproteins as co-isolates, forming part of a dynamic EV “corona” (Minimal Information for Studies of Extracellular Vesicles–MISEV 2023),^7^ we acknowledge that both EVs and lipoproteins are detectable by the NTA. Nevertheless, the presence of lipoproteins should have a minimal effect on the specificity of our EV- Trop2 ELISA since Trop2 protein is localized on the EV surface.

To further ensure the purity of the isolated EVs, we conducted Western Blot analysis by following MISEV 2018^51^ and 2023^7^ guidelines. This analysis revealed the presence of several protein markers for EVs, including the membrane-bound tetraspanin CD81 and the cytoplasmic protein TSG101, across all sample groups (**Figure 2B**). Importantly, the absence of endoplasmic reticulum (ER)-derived protein contaminants, as demonstrated by the negative expression of Calnexin, confirmed the effectiveness of our method in isolating serum-derived EVs with minimal ER protein impurities.

### Optimization of EV-Trop2 ELISA using an *in vitro* cell culture model

We utilized the EVs isolated from the DU145 prostate cancer cell line to optimize our EV-Trop2 ELISA protocol (**Supplementary Figure S2**). DU145 is a well-studied metastatic prostate cancer cell line derived from brain^52^ and has been shown to have elevated cellular Trop2 level.^38^ Subsequently, we employed our sandwich ELISA method to detect EV-Trop2, as depicted in **Supplementary Figure S2A**. We quantified the number of EVs (2.19 x 10^6^ ± 8.28 x 10^4^ particles / mL culture media / 1 million cell) and assessed their size distribution via NTA (**Supplementary Figure S2B**). Furthermore, we spiked a healthy human plasma sample (purchased de-identified samples) with the EVs isolated from DU145 cell line to evaluate the assay’s specificity and determine the minimum volume of EV sample required for the development of EV-Trop2 assay (**Supplementary Figure S2C**). Using a known DU145 EV concentration of 1.9 x 10^8^ particles/µL (as calculated from **Supplementary Figure S2B**), the assay’s linearity was determined between 1.25 µL and 5 µL of EV sample. For our analysis of serum-derived EV clinical samples, 5 µL was used. To ensure assay consistency, we conducted internal batch-to-batch testing while using different ELISA plates to measure EV- Trop2 levels from clinical samples. Across three plates, we obtained similar trends for calibration curves, R^2^ > 0.990, demonstrating repeatability (R^2^ Plate 1 = 0.998, R^2^ Plate 2 = 0.998, R^2^ Plate 3 = 0.992) (**Supplementary Figure S2D**).

### Measurements of EV-Trop2 levels

In our discovery cohort (N = 66), EV-Trop2 levels were highly elevated in the high-risk PC group (**Figure 2C**). The differences were statistically significant in differentiating between the high-risk PC and the cancer-free groups (p < 0.0001), and the high-risk PC and the low-risk PC groups (p = 0.0015). We demonstrated that the elevated levels of EV-Trop2 could be indicative of high-risk PC. The EV-Trop2 levels were normalized to the EV number (from NTA readout) in the 5 µL EV solution used for the ELISA. This normalization step, as described in the Methods section, was performed to account for variability in the EV quantities across patients’ samples (**Supplementary Figure S3**). Furthermore, to validate our EV-Trop2 ELISA platform, we conducted Western Blot analyses on selected samples and subsequently compared the chemiluminescence intensity results with the EV-Trop2 ELISA measurements (**Supplementary Figure S4**). As the ELISA readouts indicated higher levels of EV-Trop2 (*e.g.,* in the case of high-risk PC), we consistently observed a corresponding increase in the chemiluminescence results from Western Blot, confirming the existence of Trop2 on EVs and their accessibility to the antibodies used in ELISA.

To compare EV-Trop2 levels across cancer-free samples, low-risk and high-risk prostate cancer samples, box plots were used to visually show the distribution of the data (**Figure 2D**). Specifically, the median (Q2) of the high-risk PC group (2.603 x 10^-9^ pg/particle; 95% CI 1.893–3.589) exhibited a significant elevation compared to both of the cancer-free (0.426 x 10^-9^ pg/particle; 95% CI 0.234–0.808) and low-risk PC (0.752 x 10^-9^ pg/particle; 95% CI 0.453–1.844) groups. It is important to clarify that in the case of EV-Trop2 levels, a few negative values were observed as a result of the linear fitting of the calibration curve of the Trop2 standards, rather than being negative absorbance readings (**Supplementary Figures S2D and S3A–B**). When comparing the low-risk and high-risk PC groups, there were some overlaps; however, as we will demonstrate later, selecting the appropriate threshold for EV-Trop2 improved the classification of high-risk PC patients (**Figure 3**).

**Figure 3.**
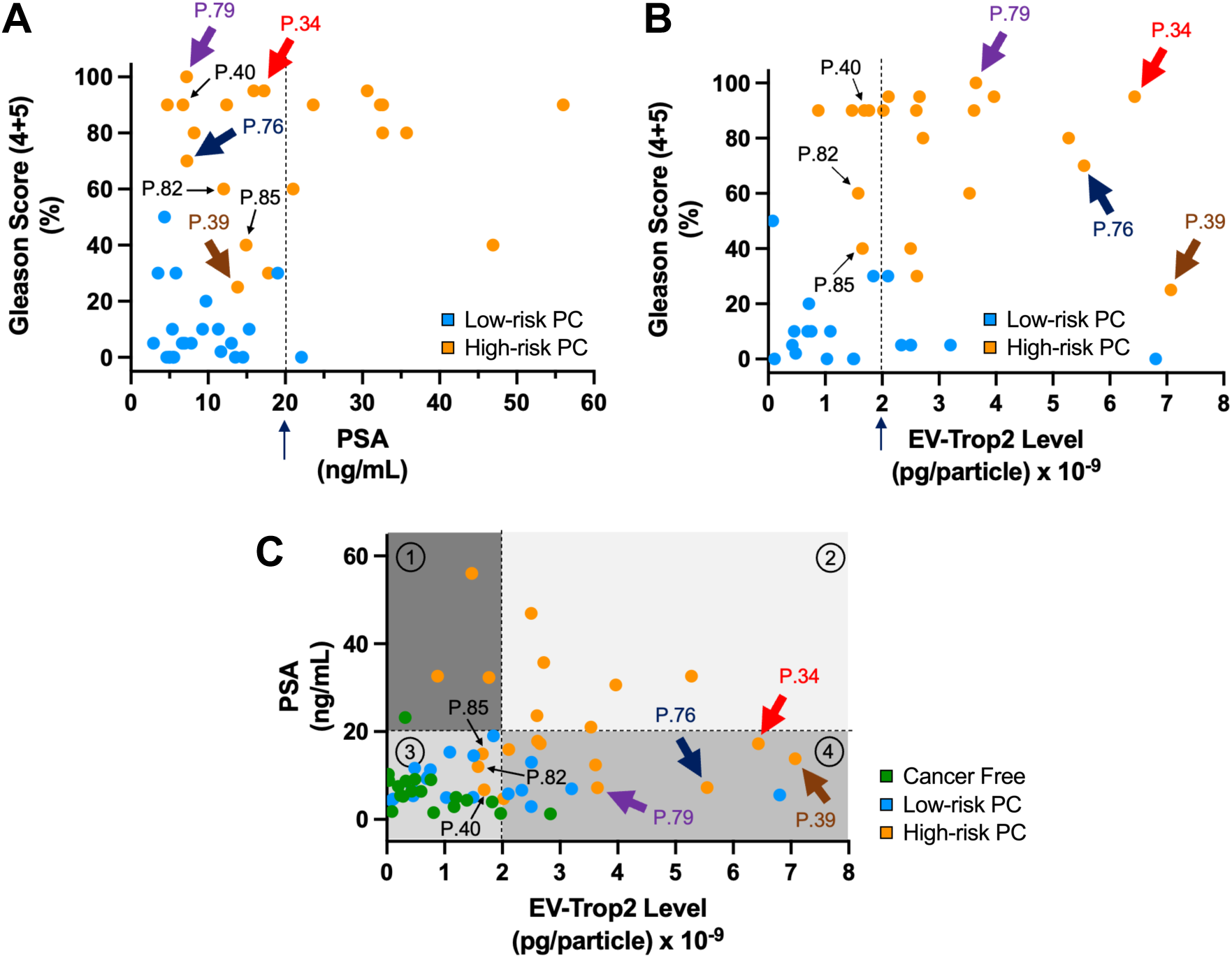
Establishing EV-Trop2 threshold level relevant to detecting high-risk PC. **(A)** Correlation between %Gleason score (Grade 4+5) and PSA levels. **(B)** Correlation between %Gleason score (Grade 4+5) and EV-Trop2 levels. **(C)** Correlation between PSA and EV-Trop2 levels. Large arrows indicate instances of high-risk PC patients with low PSA levels but elevated EV-Trop2 levels, highlighting the potential of combined analysis for improved risk stratification. Small arrows represent cases of high-risk PC patients that were not identified by either the PSA or EV-Trop2 test.

### Determining a threshold of EV-Trop2 level relevant to detecting high-risk PC patients

Prostate cancer poses a distinct clinical challenge, given the imperative to distinguish patients who require treatment (*i.e.,* high-risk prostate cancer) from those who can be managed through active surveillance (*i.e.,* low-risk prostate cancer) in a minimally invasive fashion.^53–55^ A primary motivation for this focus is the inherent limitation of the PSA test to discriminate between benign conditions and prostate cancer^56^ because PSA is expressed by both normal and malignant prostate tissues.^57^ These findings underscore the limitation of routine PSA testing, as it may potentially lead to an overdiagnosis of healthy individuals as patients, an overprescribed solid biopsy that may still not adequately capture tumor, or overtreatment of low-risk prostate cancer cases.^58,59^

According to the guidelines by the American Urological Association (AUA)^60^ and Prostate Cancer Foundation (PCF),^61^ a PSA level exceeding or equal to 20 ng/mL may indicate high-risk prostate cancer, whereas PSA levels <10 ng/mL are categorized as low risk and those 10–<20 ng/mL as intermediate risk (**Supplementary Figure S5**). Typically, patients falling into these categories are referred for pathological evaluation of their tissue biopsies to obtain the Gleason scores. While only looking at PSA levels, our study cohort revealed that 12 high-risk PC patients would have been otherwise misclassified as low-risk PC or cancer-free because they had exhibited PSA levels below 20 ng/mL (**Figure 3A**). Intriguingly, by establishing a threshold for EV-Trop2 at 2 x 10^-9^ pg/particle (> 90^th^ percentile of cancer-free and > 75^th^ percentile of low-risk PC; **Figure 2D**), fewer high-risk PC patients (now only 6, 50% reduction) were misclassified as cancer-free or low-risk PC (**Figure 3B**).

Incorporating EV-Trop2 alongside PSA improved PC risk classification, especially when distinguishing high-risk PC patients from low-risk PC patients and cancer-free individuals. This improvement was evident in the identification of high-risk PC patients exhibiting high EV-Trop2 levels but low PSA levels. As an example, four high-risk PC patients, namely Patient 34, Patient 39, Patient 76, and Patient 79, had PSA levels below 20 ng/mL, suggesting they could have been placed in the low-risk or cancer-free in the absence of a tissue biopsy. Such misclassification could have serious consequences, including the silent progression of the focal cancer to the regional or distant metastasis, potentially limiting treatment options and increasing the morbidity and mortality risk. In contrast, the EV-Trop2 levels of these patients exceeded the recommended assay threshold (> 2 x 10^-9^ pg/particle), indicating that EV-Trop2 could serve as a complementary test to PSA.

### EV-Trop2 as a potential complementary blood-based marker to PSA test in detecting high-risk PC patients

To further explore the complementary potential of EV-Trop2 alongside PSA testing, we compared EV-Trop2 and PSA levels across the three sample groups. As shown in **Figure 3C**, the results reveal four quadrants for classifying prostate cancer patients. These quadrants are as follows: 1) low EV-Trop2, high PSA; 2) high EV- Trop2, high PSA; 3) low EV-Trop2, low PSA; 4) high EV-Trop2, low PSA. The majority of high-risk PC patients fall within Quadrants 2 and 4, signifying that high EV-Trop2 levels are significant in identifying high- risk PC patients regardless of PSA levels. Quadrant 4 becomes particularly important, especially when considering the four high-risk PC patients mentioned earlier, who might have been misclassified due their low PSA levels. This highlights that high EV-Trop2 levels, even in the presence of low PSA levels, could be indicative of high-risk PC. However, there are a few misclassified low-risk PC and cancer-free cases in Quadrant 4 that might have been caused potentially by other contributing factors related to the individual patients. Hence, it warrants the requirement for larger cohort studies to further explore the utility of our combined approach. Moreover, there are cases where neither the EV-Trop2 test nor the PSA test accurately distinguished between low-risk PC and cancer-free groups. This is referred to as Quadrant 3 in the analysis. In addition, three high-risk PC patients (*e.g.,* Patient 40, Patient 82, and Patient 85), also falling into Quadrant 3, were misclassified as low-risk PC or cancer-free groups. Distinguishing low-risk PC and cancer-free patients proved more challenging in this study cohort due to the elevated PSA levels in the cancer-free group (> 5 ng/mL) compared to the recommended guidelines (< 4 ng/mL).^62,63^ On the other hand, there is an exception where the PSA test identified three high-risk PC patients that were not detected by the EV-Trop2 test (Quadrant 1). This might be due to the selection of Trop2 on EVs, which specifically targets certain tumor-associated EV populations but might not capture all types of EVs relevant to prostate cancer. While we show Trop2 is associated with tumor EVs, it is also worth noting that other non-tumor-derived EVs are also present in the serum. Nonetheless, our findings suggest that EV-Trop2 could be employed as a valuable signature in blood to complement PSA and enhance the classification of the high-risk PC group, as shown in Quadrant 2 and Quadrant 4.

### Combining EV-Trop2 and PSA enhances the risk group stratification in machine learning model of prostate cancer patients

We undertook a comprehensive multivariate analysis to develop models and training strategies that leverage EV-Trop2 and PSA to effectively stratify prostate cancer patients and healthy individuals. This involved a comparative performance measure implementing various machine learning algorithms,^64^ including k-Nearest Neighbors (kNN), Logistic Regression, Naïve Bayes, AdaBoost, Neural Network, Stochastic Gradient Descent (SGD), Random Forest, Gradient Boosting, and Support Vector Machines (SVM). We assessed their performance by examining receiver operating characteristic (ROC) curves (**Supplementary Figure S6**) and ranking them based on the F1 score, a crucial metric that balances precision (specificity) and recall (sensitivity). A higher F1 score indicates superior algorithm performance. Our goal was to minimize false negatives (increasing Recall) and false positives (increasing Precision). As a result, the Neural Network emerged as the top-performing model overall for risk classification (<u>PSA</u>: F1 score = 0.487, Precision = 0.492, Recall = 0.485; <u>EV-Trop2 + PSA</u>: F1 score = 0.607, Precision = 0.611, Recall = 0.606). Neural Network consists of interconnected layers of nodes (or neurons) that can learn from complex patterns through a process of adjusting weights based on input data.^65^ They are particularly effective with high-dimensional data and can capture non-linear relationships. To address the issue of potential overfitting, we fine-tuned the regularization parameters within a range of 0.0001 to 0.09. Within this range, the model’s performance remained stable using a leave-one-out cross validation (LOOCV) strategy. However, increasing the regularization strength beyond this threshold may overly constrain the model and could lead to underfitting, which was seen by the decrease in the performance metrics for specific groups, such as worse outcome metrics in low-risk PC (accuracy decreases by 1.5%, PPV decreases by 2.6%) and cancer free (accuracy decreases by 1.5%, PPV decreases by 2.3%). This finding suggests that the chosen regularization range (0.0001–0.09) was optimal for this dataset, although further validation in a larger cohort is necessary.

When applying the Neural Network algorithm to classify the three patient groups, the combination of EV- Trop2 and PSA showed enhanced sensitivity, specificity, and accuracy compared to using PSA alone (**Figure 4A and Figure 4B**). For instance, PSA alone accurately classified 78.8% of high-risk PC patients, whereas when EV-Trop2 and PSA were used together, this accuracy increased to 84.8%. The positive predictive values (PPV) improved across all three groups (greater than or equal to 10%). Moreover, we statistically compared the performance of EV-Trop2 + PSA against PSA alone using McNemar’s test,^66^ yielding a p-value of 0.16. Although this result suggests that the observed performance improvement is not statistically significant in risk classification, likely due to the limited sample size (N = 66), our findings indicate that the addition of EV- Trop2 enhances the model’s performance metrics (*e.g.,* sensitivity, specificity, and accuracy, PPV, and NPV), as shown in **Figure 4A and Figure 4B**. This is also evident from the ability of EV-Trop2 levels to differentiate high-risk PC from low-risk PC (p = 0.0015) and cancer-free samples (p < 0.0001) (**Figure 2C**). With validation in a larger cohort, EV-Trop2 could be a valuable complementary test to PSA screening for prostate cancer diagnosis before ordering a tissue biopsy. This strategy has the potential to alleviate patient distress and reduce unnecessary biopsy risks.

**Figure 4.**
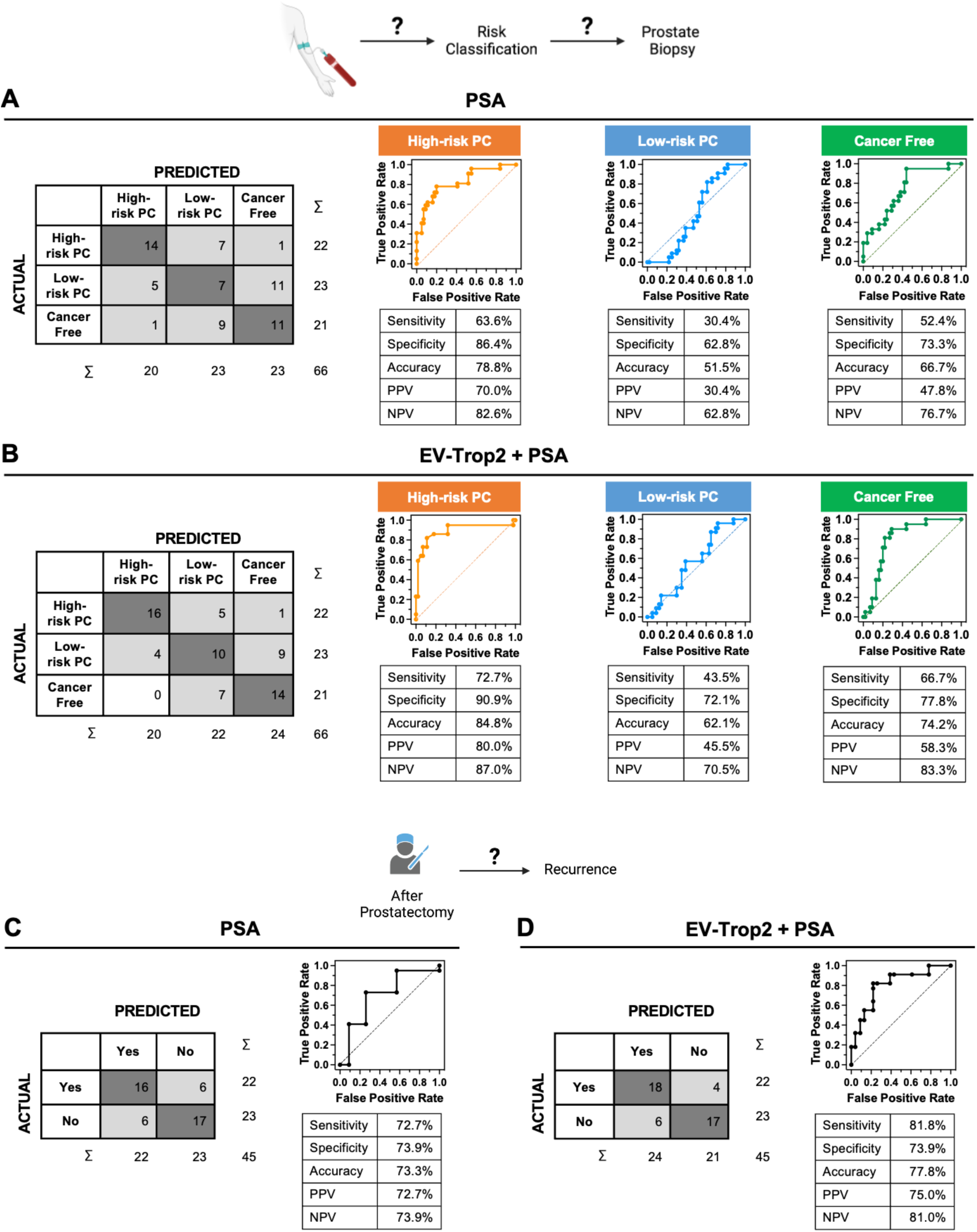
Machine learning model for risk stratification and recurrence prediction using EV-Trop2 and PSA levels. The confusion matrix and ROC curves using Neural Network algorithm show the predicted and actual classifications for each sample group, comparing **(A)** the use of PSA alone versus **(B)** the integration of EV-Trop2 as a complementary test to PSA for patient risk stratification. The recurrence prediction classifier after prostate removal surgery (prostatectomy) is compared using **(C)** PSA alone versus **(D)** the combination of PSA and EV-Trop2 using Naïve Bayes algorithm. PPV = positive predictive value; NPV = negative predictive value.

Additionally, we performed an error analysis to identify the features contributing to misclassification in our machine learning models (**Supplementary Figure S7**). This analysis focused specifically on the misclassifications between (i) low-risk PC (LR) and cancer-free (CF) groups, as well as between (ii) high-risk PC (HR) and low-risk PC (LR) groups. The confusion matrix revealed that these two comparisons exhibited the highest misclassification rates, while it was the lowest for the classification between HR and CF. In our error analysis, we observed distinct patterns in both the PSA-only and the EV-Trop2 + PSA models. For the PSA-only model (**Supplementary Figure S7A**), misclassifications between HR and LR categories predominantly occurred when PSA levels were below 14 ng/mL, leading to HR samples being incorrectly classified as LR. Conversely, LR samples were misclassified as HR when PSA levels exceeded 14 ng/mL, highlighting the limitations of PSA alone, especially since clinical guidelines suggest HR is indicated by PSA levels above 20 ng/mL.^60,61^ Additionally, CF samples were misclassified as LR when PSA levels surpassed 6 ng/mL, while LR samples were misclassified as CF when PSA levels fell below 7 ng/mL, indicating frequent misclassifications in this borderline range (PSA > 4 ng/mL).^60,61^ Misclassifications between HR and CF were also noted, particularly when PSA levels were around 4 ng/mL, which is significantly lower than the clinical HR threshold (PSA > 20 ng/mL). In contrast, the EV-Trop2 + PSA model demonstrated improved classification accuracy (**Supplementary Figure S7B**), including 7 HR misclassifications as LR (using PSA only) down to 4 HR misclassification as LR (using EV-Trop2 + PSA). Misclassifications of HR as LR occurred when EV-Trop2 levels were below 2 x 10^-9^ pg/particle, while LR samples were misclassified as HR when EV- Trop2 levels exceeded this threshold. Notably, PSA levels did not show a clear trend in this model, suggesting that EV-Trop2 might serve as a potential reliable adjunct marker than PSA alone. Meanwhile, misclassifications between LR and CF were observed when EV-Trop2 levels were below 2–3 x 10^-9^ pg/particle, with CF samples misclassified as LR when PSA levels were higher than 6 ng/mL. Lastly, HR samples were misclassified as CF when PSA levels were around 8 ng/mL and EV-Trop2 levels were significantly lower than 2 x 10^-9^ pg/particle. This finding is consistent with our discussion earlier and is further illustrated in **Figure 3C**. It shows the limitations of setting the 2 x 10^-9^ pg/particle EV-Trop2 threshold. This threshold led to the exclusion of a few high-risk PC cases, which points to the need for validating this assay in larger cohorts.

### Potential use of combining EV-Trop2 and PSA in predicting recurrence after surgery using machine learning model of prostate cancer patients

Further, our study demonstrates a correlation between EV-Trop2 level and Gleason score (a key indicator of prostate cancer aggressiveness and prognostication) (**Figure 3B**); hence, elevated EV-Trop2 levels in patients with localized disease could have far-reaching implications such as: i) guiding the decision to perform a prostate biopsy, potentially helping oncologists to identify patients for active surveillance monitoring (watchful waiting) (**Figures 4A–B**), and ii) aiding post-surgery decisions regarding follow-up to monitor recurrence (**Figures 4C–D**). Building on the observed correlation between EV-Trop2 level and Gleason score, we evaluated the predictive value of EV-Trop2 for recurrence following surgical treatment (prostatectomy) using the Naïve Bayes algorithm (one of the top performing models; **Supplementary S8;** <u>PSA</u>: F1 score = 0.733, Precision = 0.733, Recall = 0.733; <u>EV-Trop2 + PSA</u>: F1 score = 0.778, Precision = 0.780, Recall = 0.778). Our findings indicate improved sensitivity and accuracy when combining EV-Trop2 and PSA compared to using PSA alone (**Figure 4C and Figure 4D**). When considering PSA alone, the model predicted recurrence in 73.3% of cases. However, when EV-Trop2 and PSA were combined, this accuracy increased to 77.8%. The PPV rose from 72.7% to 75.0%, while the NPV increased from 73.9% to 81.0%. Although these results show that our combined EV-Trop2 and PSA machine learning model presents potential applications in predicting recurrence at early stages when compared to just using PSA alone (p = 0.49, McNemar’s test^66^), we acknowledge that expanding the size of the cohort could be useful to improve classification accuracy and boost the predictive power of the recurrence model. This current study, centered on EV-Trop2 and its modeling framework, serves as an initial demonstration of possible analyses and the potential of the combined biomarkers (EV-Trop2 + PSA) in predicting recurrence. We anticipate building on this work in future studies to identify the most clinically useful model.

Our machine learning results suggest that the combination of EV-Trop2 and PSA has potential applications in identifying clinically significant high-risk PC patients and in early-state recurrence prediction, increasing performance metrics compared to PSA alone. This combined approach could potentially serve as an adjunct blood-based test alongside tissue-based genetic tests^67^ (*e.g.,* Decipher, Oncotype Dx Prostate, Prolaris), offering a minimally invasive approach for risk stratification and recurrence monitoring. Meanwhile, the ExoDx Prostate Test, a non-invasive urine test, measures exosomal RNA levels of three biomarkers (ERG, PCA3, and SPDEF) to assess the risk of high-grade prostate cancer, primarily for patients with PSA levels between 2-10 ng/mL who are considering an initial biopsy.^68,69^ In contrast, our assay uses a simple ELISA to detect Trop2 protein on EVs, making EV-Trop2 as a potential blood-based adjunct test to PSA. Our approach aims to improve risk stratification, particularly to identify high-risk prostate cancer cases, and to support recurrence prediction after prostate surgery. Both ExoDx RNA test and our EV-Trop2 ELISA aim to improve diagnostic accuracy to potentially reduce biopsy frequency and patient distress, but our study also highlights EV-Trop2’s potential utility in early recurrence prediction, offering an additional non-invasive option for risk monitoring and assessment.

### Outlook

In this small cohort study, blood-based EV-Trop2 is shown to be a complementary and minimally invasive test that can improve the patient risk classification, especially in high-risk PC and cancer-free groups, when combined with PSA, that should be validated in larger cohorts. Once confirming the analytical validity of our EV-Trop2 isolation and quantification workflow in a larger cohort, we can assess its full clinical validity and utility. The overall goal is to use our blood-based EV-Trop2 as a liquid biopsy tool to improve stratification of high-risk PC, low-risk PC, and cancer-free groups, which could significantly impact treatment decisions, such as surveillance, radiation, or prostatectomy, as well as predict recurrence at early stages.

Our EV-Trop2 isolation and quantification tool is tailored to benefit individuals at high risk of prostate cancer. This includes individuals who may be undergoing regular prostate cancer screening, those with suspected prostate cancer based on clinical indications, or individuals already diagnosed with prostate cancer who require continuous monitoring and assessment of their disease progression or treatment response. Our blood-based EV-Trop2 liquid biopsy tool is a valuable complementary tool for improving early detection, risk stratification, and prostate cancer management. It can be incorporated into various medical practices to enhance these aspects of prostate cancer care. For example, in the assessment of Trop2 levels in blood for therapy monitoring^40,70^ upon successful completion of phase II clinical trial of Trop2-targeted therapy (*e.g.,* Trodelvy®) for metastatic castration-resistant prostate cancer (NCT03725761). Currently, Trodelvy® holds FDA approval for the treatment of triple-negative breast cancer,^71^ hormone-positive breast cancer,^72,73^ and urothelial cancer.^74^ However, there is a lack of rapid and minimally invasive tools to predict and monitor responses to Trop2- targeted therapies. Hence, our EV-Trop2 tool could potentially offer a modality for therapy monitoring, extending its potential utility beyond prostate cancer to encompass breast cancer and urothelial cancer as well.

### Study limitations

First, the relatively small sample size might limit the generalizability of our findings. Although we provide initial evidence that EV-Trop2 is a promising minimally invasive biomarker for differentiating between high- risk and low-risk PC, further validation in larger and more diverse patient cohorts, including intermediate-risk cases, is necessary to strengthen these findings. Moreover, the serum samples used in this study were all collected before mp-MRI was used routinely in clinical practice, and therefore PI-RADS scores are not available for our patient cohort. Our goal was to test whether the measurement of EV-Trop2 as a single test demonstrated any signal in detecting clinically significant prostate cancer (*e.g.,* high-risk PC), and our findings suggest that it is a promising assay (**Figure 2C**). Assessing whether EV-Trop2 adds value to MRI in clinical practice will be a critical step for its translation. However, it is worth noting that the value of many available serum or urine biomarkers (*e.g.,* 4K score, PHI, PCA3, MIPS, Stockholm3) in complementing MRI also remains unclear, and this is currently an active area of research, with reference sets being collected by the Early Detection Research Network (EDRN) and other research entities. Emerging evidence suggests that some of these tests, such as 4K score^75^ and Stockholm 3^76^, can be used to triage patients who do not need mp-MRI or biopsy. Moving forward, we plan to validate our findings in contemporary clinical cohort where mp-MRI data are available, addressing this knowledge gap.

Additionally, while we examined EV-Trop2’s diagnostic potential, its utility in monitoring treatment response and disease progression has not yet been explored. Longitudinal studies with extended follow-up periods in prostate cancer patients are essential to establish the relationship between EV-Trop2 levels and clinically meaningful outcomes, such as progression, recurrence, and survival.

In addition, we acknowledge that without a larger sample size, it is challenging to develop a robust prediction model. Adequate sample size is essential for machine learning-based modeling to achieve a more comprehensive understanding of the biological and molecular mechanisms underlying prostate cancer, particularly in the context of risk classification and recurrence prediction. Future studies involving larger cohorts could further validate the potential benefit of incorporating EV-Trop2 and PSA into the machine learning model, with the aim of enhancing classification accuracy and improve the predictive power of the recurrence model. This current study serves as a preliminary demonstration of the analytical approaches that can be applied and highlights the potential of the combined signature (EV-Trop2 + PSA) in recurrence prediction. Expanding this research work in future studies would be the next critical step to validate our findings and develop an optimized model with potential clinical utility.

## CONCLUSION

The isolation and characterization of EVs, along with the quantification of Trop2 on the surface of EVs (*i.e.,* EV-Trop2) in serum samples of prostate cancer patients, can differentiate aggressiveness of prostate cancer (*e.g.,* high-risk PC from low-risk PC and cancer-free groups) and enhance the classifier model for risk stratification when combined with PSA through multivariate analysis. This integrated approach has the potential to reduce misclassification of high-risk cases. Furthermore, the combination of EV-Trop2 and PSA can enhance the classifier model for recurrence prediction after prostatectomy. Our ELISA-based EV liquid biopsy assay is easily accessible and offers the opportunity for validation in larger clinical cohorts, leading to more accurate diagnoses and personalized treatment approaches. Therefore, this tool could potentially serve as a complementary liquid biopsy tool to PSA testing to identify patients at risk for aggressive prostate cancer and predict the risk of recurrence, as well as an adjunct blood-based test to the genetic test on prostate tissues.

Expanding beyond its clinical application in assessing the aggressiveness risk of prostate cancer, this tool for EV-Trop2 holds remarkable promise across multiple epithelial cancers. It can be used in monitoring treatment responses and guiding therapeutic decisions, particularly with the emergence of Trop2-targeted therapies.

## METHODS

### Optimization of the Trop2 ELISA protocol using an *in vitro* cell culture model

#### Cell culture

DU145 cell line was purchased from ATCC (ATCC, HTB-81). The cell line was cultured in a complete growth media of RPMI-1640 with L-glutamine (Corning, 10-040-CV), as well as 10% Fetal Bovine Serum (FBS) (Cytiva, SH30910) and penicillin-streptomycin antibiotics (Cytiva, SV30010). After seeding the cells and about 48 h in the complete growth media (up to ∼80–90% confluency; ∼8–9 million cells/plate), the cells were depleted from the complete media, rinsed with phosphate-buffered saline (PBS) (Cytiva, SH30256), and replenished using media with only penicillin-streptomycin (*i.e.,* serum-deprived media). After they were depleted for approximately 48 h, the cells were detached and collected from the plates using 0.25% Trypsin (Cytiva, SH30042). The collected cells were centrifuged at 500 x g for 5 min, and then replated. This process was repeated in order to have the cells passaged again.

#### Culture media collection for EV isolation

EVs were isolated by collecting the serum-free media after the 48 h of depletion period of the culture. The collected media (∼144mL; ∼12mL/plate) was centrifuged (Thermo Scientific Sorvall ST 16R, TX-400 Rotor) at 500 x g for 5 min. The EV media was poured into a new tube to remove any cells, and then continued in the centrifuge (Thermo Scientific Sorvall ST 16R, TX-400 Rotor) at 2,000 x g for 20 min. After the EV media had been centrifuged for 20 min, the supernatant was collected. The supernatant was then kept in the freezer at - 80°C. Once it was ready to be isolated, the supernatant was thawed back to room temperature. Using four tubes at a time, they were put for ultracentrifugation (Thermo Scientific WX Ultra Series, Surespin 630 Rotor) at 100,000 x g for approximately 3 h. The supernatant was removed, and the remaining bottom layer was collected into one tube and filtered through a 0.22-µm syringe filter before isolation using ExoTIC devices, as described in the next section below.

### Patient samples

We evaluated 66 samples from the Stanford Urology Tissue and Serum Bank. Patient groups are categorized as follows:

- Cancer Free: 21 men with biopsy-confirmed absence of prostate cancer
- Low-risk Prostate Cancer: 23 men with prostate cancer who were cured by radical prostatectomy (no biochemical recurrence for at least 5 years after surgery)
- High-risk Prostate Cancer: 22 men with prostate cancer men who recurred after radical prostatectomy The patient information is summarized in Supplementary Table S1, comprising pre-operative PSA level, and %Gleason score (Grade 4+5). We have broken down samples by the percentage of Gleason patterns 4 or 5 because data from our institution demonstrates a strong correlation with adverse outcomes (*e.g.,* recurrence after surgery).^49^ The resource has been continuously reviewed and approved by the Institutional Review Board of Stanford University. The samples have been used in compliance with the IRB approved protocol #59490. All experiments were carried out in accordance with the relevant ethical regulations of Stanford University.

### Extracellular vesicle (EV) isolation and analysis

EVs were isolated using the Exosome Total Isolation Chip (ExoTIC), following the methodology outlined in our previous studies.^42,44–47^ The ExoTIC device comprised a stack of membranes, including a 50-nm polycarbonate nanoporous low protein binding filter membrane (Whatman Nucleopore), a 200-nm PES support membrane (Sterlitech), and a thick cellulose paper pad (MilliporeSigma). The isolation process began by diluting 100 µL of serum sample in PBS and then passing it through a 0.22-µm PES syringe filter (Argos Technologies). Unlike the *in vitro* samples, the serum samples did not undergo prior ultracentrifugation at 100,000 x g before ExoTIC processing. The filtered sample was then introduced into the ExoTIC device at a constant rate of 5 mL/h using a syringe pump (PHD Ultra, Harvard Apparatus, USA). Within the ExoTIC device, EVs of an approximate size range between 50 and 220 nm were retained in the isolation chamber, located in front of the filter membrane. In contrast, smaller molecules such as free-floating nucleic acids and proteins, presumed to be smaller than 50 nm, passed through the outlet. After isolating the EVs, a thorough washing step with PBS was performed to remove any remaining contaminants. Once the washing process was complete, the EV isolate was collected from the designated inlet and subjected to analysis using Nanoparticle Tracking Analysis (NTA). For the NTA procedure, the EV sample was diluted with PBS, following the instructions provided in the manufacturer’s manual for the Nanosight NS300 instrument (Malvern, UK). The specific NTA settings used were as follows: camera level = 14, capture script = 3 x 30 s, and detection threshold = 5.

### Western Blot

EV sample from each patient group was lysed using an equal part of RIPA buffer (Thermo Scientific, 89901). The protein was then reduced using Laemmli buffer (Bio-Rad, 1610747) and 2-mercaptoethanol (Bio-Rad, 1610710), followed by heating at 95°C for 5 min. Each sample was loaded into a Mini-PROTEAN® TGX™ Precast 4-20% PAGE protein gel (Bio-Rad, 4561094) and aligned at 50 V, followed by 80 V for 4 h. The proteins were then transferred from the gel to PVDF membrane by Trans-Blot Turbo Transfer System (Bio- Rad, 1704156) for 30 min at 1 A. Membranes were blocked in a blocking buffer (Bio-Rad, 12010020) and then incubated with primary antibodies for positive EV markers: mouse monoclonal to CD81 (Thermo Fisher Scientific, 10630D), mouse monoclonal to TSG101 (Novus Biologicals, NB200-112); negative EV marker: rabbit polyclonal to Calnexin (Cell Signaling, 2433S) in blocking buffer overnight at 4°C. Membranes were washed in TBST and then incubated with goat anti-rabbit IgG HRP (Cell Signaling, 7074P2) or goat anti- mouse IgG HRP (Invitrogen, 31432) in blocking buffer for 2 h at room temperature. Membranes were washed in TBST, and the PVDF membranes were then exposed to chemiluminescent substrates (Advansta, K-12043- D10) and illuminated using IVIS® Spectrum imaging system (Perkin Elmer). Chemiluminescent signals were quantified using the freely available ImageJ software.^77^ To do this, a specific region of interest (ROI) was delineated, and the pixel density was recorded for each sample. We then calculated the inverted pixel density to quantify the chemiluminescent signals. To eliminate background noise, all values were corrected by subtracting the background signals.

### Cryogenic Electron Microscopy (cryo-EM)

EVs were imaged on cryo-EM and prepared on C-flat EM grids (Electron Microscopy Sciences, USA). The grids were first prepared by glow-discharge (30 s, 25 mA) in a Pelco EasiGlow system. The glow-discharged grid was place in the humidity chamber of the instrument (Vitrobot Mark IV, FEI, USA) at 100% humidity and 4 °C. An aliquot (3 µL) of the EVs sample was then applied to the carbon side of EM grids and subsequently blotted for 3 s at a blot force of 2 and plunge-frozen into the precooled liquid ethane by liquid nitrogen. The cryo-EM grids were transferred/clipped to the grid ring holder and then loaded to the autoloader system. The images were acquired using Glacios Cryo-TEM system (Thermo Fisher Scientific).

### EV-associated Trop2 (EV-Trop2) ELISA

Detection of EV-Trop2 levels was performed using a sandwich ELISA assay (**Figure 1B**). The procedure steps are discussed briefly as follows. Anti-Trop2 capture antibody (SinoBiological, 10428-MM01) was diluted 1:100 in coating buffer (0.2 M sodium bicarbonate, pH 9.4) and incubated at 4 °C overnight to coat the ELISA plate (Corning, 3690). Then, the plate was blocked with 5% BSA as a blocking buffer, incubated overnight at 4 °C to prevent non-specific binding. To determine the optimal volume of EVs for our in-house assay, we utilized EVs isolated from the DU145 cell line spiked into healthy human plasma, with varying EV volume from 1.25 µL to 5 µL. After determining the optimal volume that fell within the linear range (**Supplementary Figure S2C**), EV samples obtained from cancer-free patients, low-risk PC patients, and high-risk PC patients (5 µL each) were added to the ELISA plate and incubated for 2 h at room temperature. Next, biotinylated anti- Trop2 detection antibody (R&D Systems, BAF650), diluted 1:250 in blocking buffer, was added to the plate and incubated for 2 h at room temperature. Afterward, the plate was washed three times with PBST (PBS containing Tween-20) to remove unbound antibody. Then, streptavidin-HRP (Thermo Fisher Scientific, PI21134), diluted 1:1000 in blocking buffer, was added to the plate and incubated at room temperature for 30 min. After three washes with PBST and two washes with PBS, the signals were detected using ultra TMB- ELISA substrate (Thermo Fisher Scientific, 34028) after incubating the plate for 20 min. The reaction was stopped by adding stopping solution (Thermo Fisher Scientific, PIN600), and the absorbance was then measured three times using a plate reader (Promega, GloMax) at 450 nm. Meanwhile, a standard curve was generated using human recombinant Trop2 protein (SinoBiological, 10428-H08H-1) with a concentration range of 0–0.5 pg/µL.

### EV-Trop2 ELISA data analysis

We provided the raw absorbance values from the ELISA of each patient sample in **Supplementary Figure S3A**. The values were normalized by subtracting the absorbance of the blank control (0 pg) from each sample, using the calibration curves presented in **Supplementary Figure S2D (i)**. The normalized absorbance of the calibration curve is shown in **Supplementary Figure S2D (ii)**, for reference. Then, the corresponding EV- Trop2 levels (pg) for each sample were calculated from the normalized absorbance and are displayed as a line graph overlaid on the bar graph of normalized absorbance (**Supplementary Figure S3B**). Subsequently, we normalized the EV-Trop2 level (pg) to the concentration of EV (particles/µL), measured by NTA, and the EV volume used for ELISA (µL). The EV concentration of each sample is shown in **Supplementary Figure S3C**. The final reported EV-Trop level is in pg/particle (**Figure 2C**), calculated by

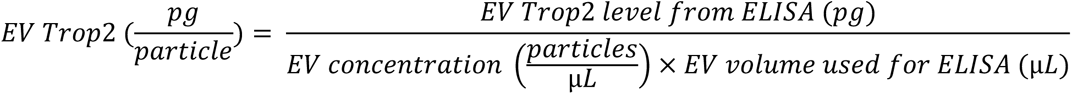

where EV volume used for ELISA was 5 µL from all serum samples.

### Data processing and visualization

Data analysis and plotting were done using GraphPad Prism version 10.1.0 for macOS (GraphPad Software, Boston, Massachusetts, USA, www.graphpad.com), Python-based Orange Data Mining software,^64^ and PlotsOfData web app.^78^ Machine learning algorithm was run using Python-based Orange Data Mining software. Schematic drawing was created with BioRender.com.

A range of machine learning algorithms, including k-Nearest Neighbors (kNN), Logistic Regression, Naïve Bayes, Neural Network, Random Forest, Support Vector Machines (SVM), Stochastic Gradient Descent (SGD), Gradient Boosting, and AdaBoost was employed to create models and training strategies for our multivariate datasets. For model and training strategy evaluation, Receiver Operating Characteristic (ROC) curves were generated for each algorithm using leave-one-out cross validation (LOOCV), allowing the performance of our classification models and training strategies to be assessed (**Supplementary Figure S6 and Supplementary Figure S8**). The F1 score, a crucial metric that measures the harmonic mean of precision and recall, was the focus of the performance evaluation. A higher F1 score signifies better overall algorithm performance. Additionally, confusion matrices were constructed for each algorithm, and their performance was compared with respect to the target patient groups.

### Parameters for machine learning algorithms

#### kNN

- Number of neighbors = 5
- Distance metric = Euclidean
- Weighting function = Uniform

#### Logistic Regression

- Regularization type = Lasso (L1)
- Strength (C) = 1

#### Naïve Bayes

- No individual settable parameters, but by default, the numeric values are discretized into four bins with equal frequency

#### Neural Network

- Neurons in hidden layers = 100
- Activation = ReLu
- Solver = Adam
- Regularization (*α*) = 0.0001–0.09
- Maximal number of iterations = 200
- Replicable training = Yes

#### Random Forest

- Number of trees = 10
- Replicable training = Yes
- Limit depth of individual trees = 5
- Do not split subsets smaller than = 5

#### Support Vector Machines (SVM)

- SVM Type = SVM
- Cost (C) = 1
- Regression loss epsilon (*ε*) = 0.1
- Kernel = RBF; g = auto
- Numerical tolerance = 0.001
- Iteration limit = 100

#### Stochastic Gradient Descent (SGD)

- Loss function classification = Hinge
- Loss function regression = Squared Loss
- Regularization = Ridge (L2)
- Regularization strength (*α*) = 0.00001
- Optimization learning rate = Constant
- Initial learning rate (*η*_0_) = 0.01
- Inverse scaling exponent (t) = 0.25
- Number of iterations = 1000
- Tolerance (stopping criterion) = 0.001
- Shuffle data after each iteration = Yes

#### Gradient Boosting

- Method = Gradient Boosting (catboost)
- Number of trees = 100
- Learning rate = 0.3
- Replicable training = Yes
- Regularization (*λ*) = 3
- Limit depth of individual trees = 6
- Fraction of feature for each tree = 1

#### AdaBoost

- Base estimator = Tree
- Number of estimators = 50
- Learning rate = 1
- Classification algorithm = SAMME.R
- Regression loss function = Linear

## SUPPLEMENTARY INFORMATION

**Supplementary Figure S1.** Nanoparticle Tracking Analysis (NTA) measurements of the serum-derived EVs.

**Supplementary Figure S2.** Optimization of EV-Trop2 ELISA.

**Supplementary Figure S3.** Analysis of EV-Trop2 ELISA data.

**Supplementary Figure S4.** Comparative analysis of serum-derived EV-Trop2 levels employing ELISA and Western Blot.

**Supplementary Figure S5.** Illustration of EV-Trop2 levels using clinical risk guidelines.

**Supplementary Figure S6.** ROC curves assessing the performance of risk group classification of nine machine learning algorithms using EV-Trop2 and PSA parameters.

**Supplementary Figure S7.** Error analysis to identify the features contributing to misclassification in our risk stratification models.

**Supplementary Figure S8.** ROC curves assessing the performance of recurrence prediction of nine machine learning algorithms using EV-Trop2 and PSA parameters.

**Supplementary Table S1.** Patient information categorized by risk groups.

**Supplementary References.**

## AUTHOR CONTRIBUTIONS

**Conceptualization:** P.D.S., M.O.O., E.C.H., T.S., U.D. **Formal analysis:** P.D.S., M.O.O., S.L., D.A. with assistance from E.D. **Methodology:** P.D.S., M.O.O., S.L., E.C.H. **Investigation:** P.D.S., M.O.O., S.L., E.D. **Resources – clinical samples:** R.N. and J.D.B. **Machine learning modeling:** P.D.S. and D.A. **Visualization:** P.D.S., with inputs and assistance from M.O.O. **Writing – original draft:** P.D.S., with assistance and contributions from M.O.O., S.L., E.D., and D.A. **Writing – review & editing:** P.D.S., M.O.O., S.L., D.A., J.D.B., T.S., U.D. **Supervision:** T.S. and U.D. **Funding acquisition:** M.O.O., E.C.H., T.S., U.D. All authors have given approval to the final version of the manuscript.

## DATA AVAILABILITY

The datasets used and analyzed in the study are available from the corresponding authors upon a reasonable request.

## ACKNOWLEGMENTS

This work was supported by the Stanford Clinical and Translational Science Award (CTSA) to Spectrum (UL1TR003142) and the SPARK at Stanford Program (through Weston-Havens Foundation). The CTSA program is led by the National Center for Advancing Translational Sciences (NCATS) at the National Institutes of Health (NIH). The content is solely the responsibility of the authors and does not necessarily represent the official views of the NIH. This work was also partly supported by NIH grant R01EB029805. P.D.S. acknowledges support from the James D. Plummer Graduate Fellowship, EDGE Doctoral Fellowship Program, Summer First Fellowship Program, Dean’s Office of the Stanford School of Engineering, Cancer Imaging & Early Detection Award, Canary-ACED Graduate Fellowship, and Stanford Bio-X Interdisciplinary Initiatives Program (IIP) Seed Grant. M.O.O. and E.C.H. acknowledge partial support from the SPARK at Stanford Program (through Weston-Havens Foundation) and NIH grant R37CA240822. P.D.S. and E.D. would like to thank the Canary CREST Program (NIH R25CA217729). T.S. is supported by the National Institutes of Health/National Cancer Institute (NCI) R37CA240822, R01CA274978, and R01CA244281, as well as the US Army Medical Research Acquisition Activity, No. GRANT13686517. S.L. is supported by the UCLA Jonsson Comprehensive Cancer Center. The authors would like to thank the SPARK community and advisors (Prof. Steve Schow and Prof. Steven Goodman) for tremendous support and valuable inputs. We would also like to thank Magda Zaoralova and Dr. Elizabeth Montabana at the Stanford University Cryo-electron Microscopy Center (cEMc) facility for support with cryo-EM data collection. The authors would also like to extend sincere appreciation to Prof. Manish Kohli (Utah), Prof. Shan X. Wang (Stanford), Timothy Somerset (Cambridge), and Daniel Jacobson (Cambridge) for their valuable comments.

