## Supplementary Information for "Circulating extracellular vesicles in serum carry Trop2 marker for prostate cancer liquid biopsy and clinical care"

### Table of Contents


1. **Supplementary Figures**
   - **S1.** Nanoparticle Tracking Analysis (NTA) measurements of the serum-derived EVs.
   - **S2.** Optimization of EV-Trop2 ELISA.
   - **S3.** Analysis of EV-Trop2 ELISA data.
   - **S4.** Comparative analysis of serum-derived EV-Trop2 levels employing ELISA and Western Blot.
   - **S5.** Illustration of EV-Trop2 levels using clinical risk guidelines.
   - **S6.** ROC curves assessing the performance of risk group classification of nine machine learning algorithms using EV-Trop2 and PSA parameters.
   - **S7.** Error analysis to identify the features contributing to misclassification in our risk stratification models.
   - **S8.** ROC curves assessing the performance of recurrence prediction of nine machine learning algorithms using EV-Trop2 and PSA parameters.

2. **Supplementary Tables**
   - **S1.** Patient information categorized by risk groups.

3. **Supplementary References**

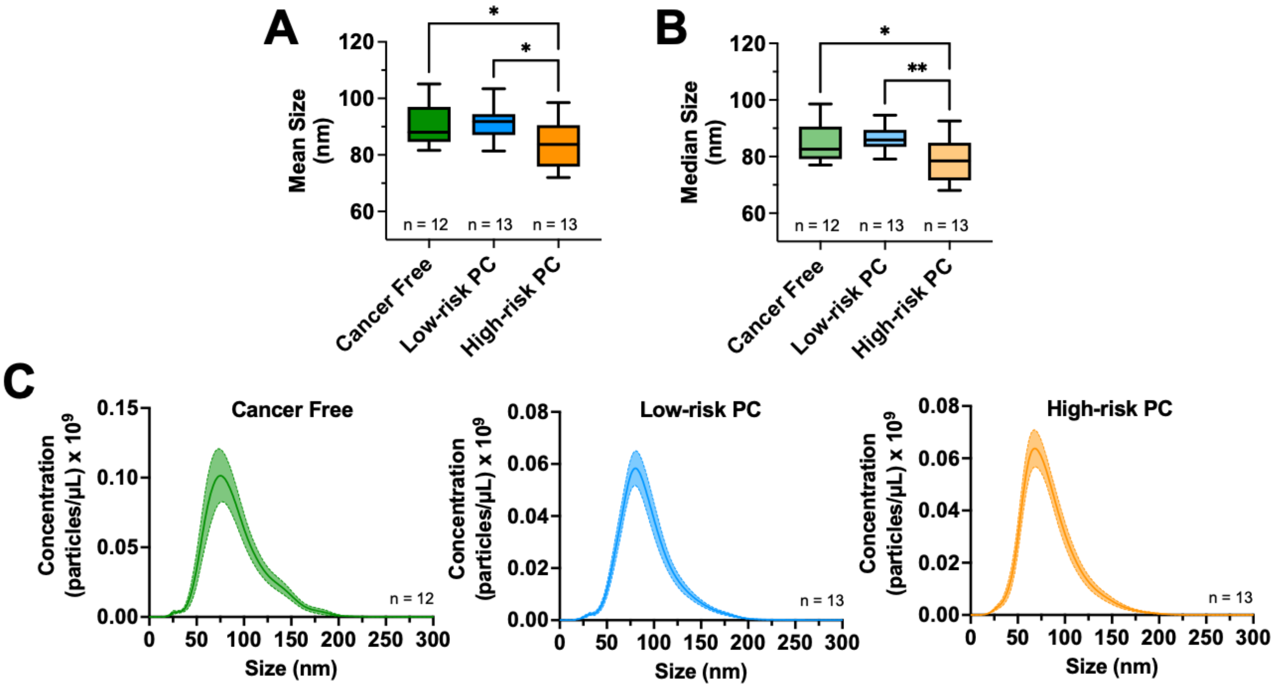


**Supplementary Figure S1. Nanoparticle Tracking Analysis (NTA) measurements of the serum-derived EVs. (A)** Mean size and **(B)** median size of the EVs from the three sample groups (cancer free, low-risk PC, and high-risk PC). Statistical analysis: One-way ANOVA with ordinary ANOVA's multiple comparison test; * $p \leq 0.05$, ** $p \leq 0.01$. **(C)** Size distributions of the EVs from the three sample groups. Shaded areas are error bands.

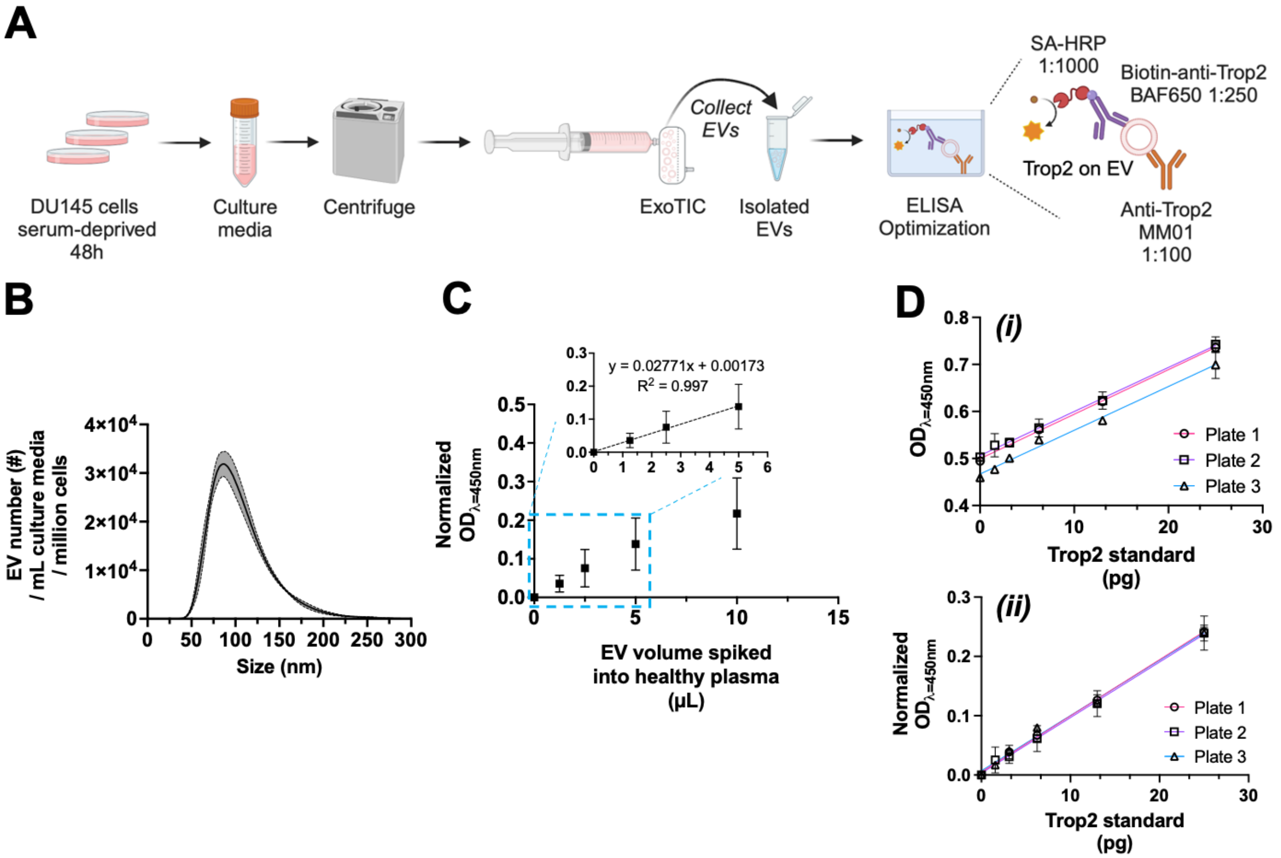


**Supplementary Figure S2. Optimization of EV-Trop2 ELISA. (A)** Schematics illustrating the EV-Trop2 isolation and detection using EVs from DU145 cell culture. The conditioned culture media are subjected to ultracentrifugation, followed by isolation using our device, ExoTIC. The collected EVs are then used in ELISA for the detection of EV-Trop2. Figure was created with BioRender.com. **(B)** Size and concentration distribution of the collected EVs measured using Nanoparticle Tracking Analysis (NTA). Shaded area is error band. **(C)** Optimization of EV sample volume for ELISA by spiking the EVs into healthy human plasma and measuring $OD_{450nm}$; the linear range is observed at 1.25–5 µL of EV sample volume. Error bars are standard deviations. **(D)** Batch-to-batch study comparing the Trop2 standards used in different ELISA plates for separate EV-Trop2 measurements from clinical serum samples ($R^2 > 0.990$). Error bars are standard deviations.

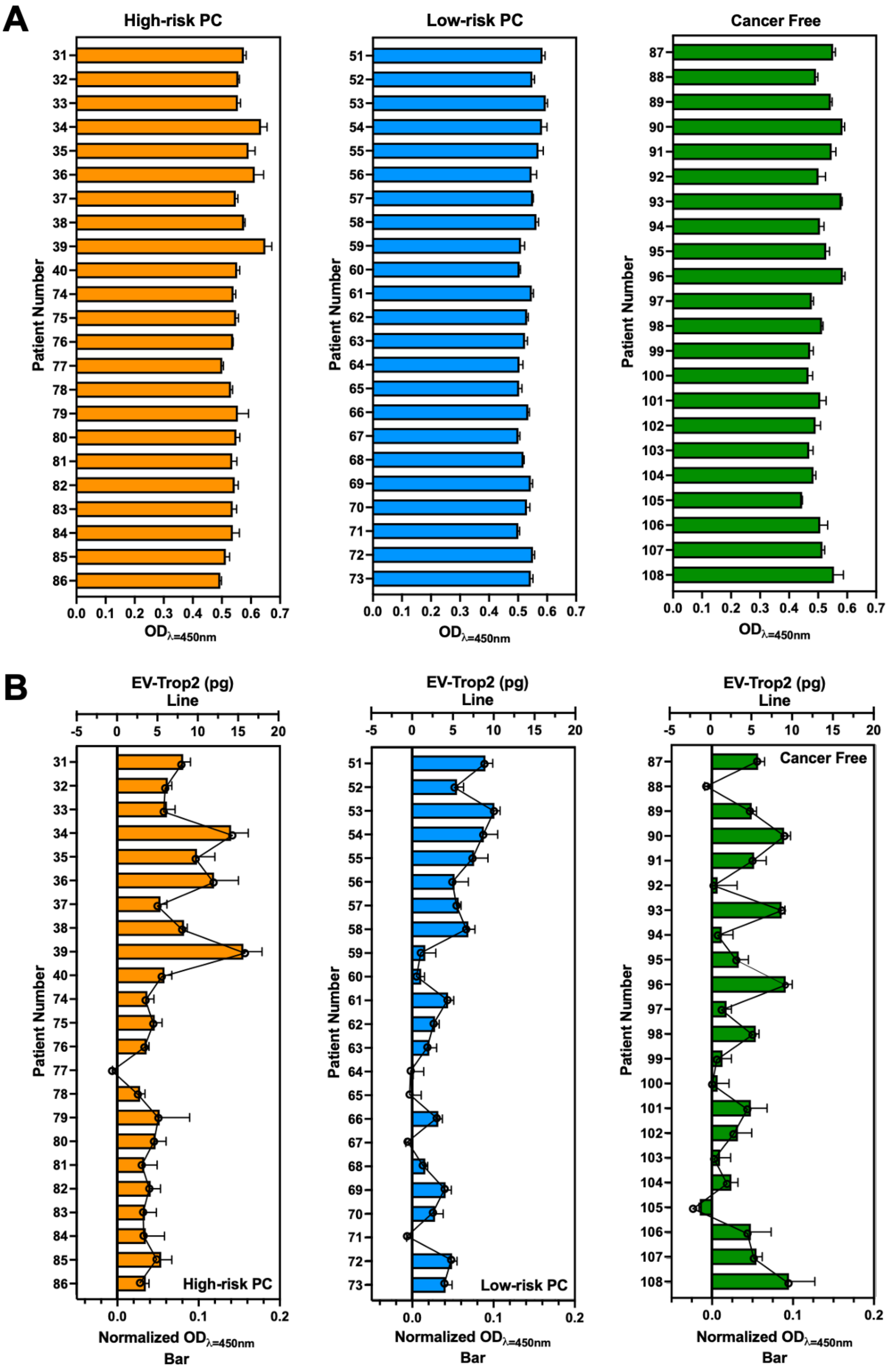
A
High-risk PC
Low-risk PC
Cancer Free
Patient Number
OD λ=450nm
B
EV-Trop2 (pg)
Line
Normalized OD λ=450nm
Bar
High-risk PC
Low-risk PC
Cancer Free

1

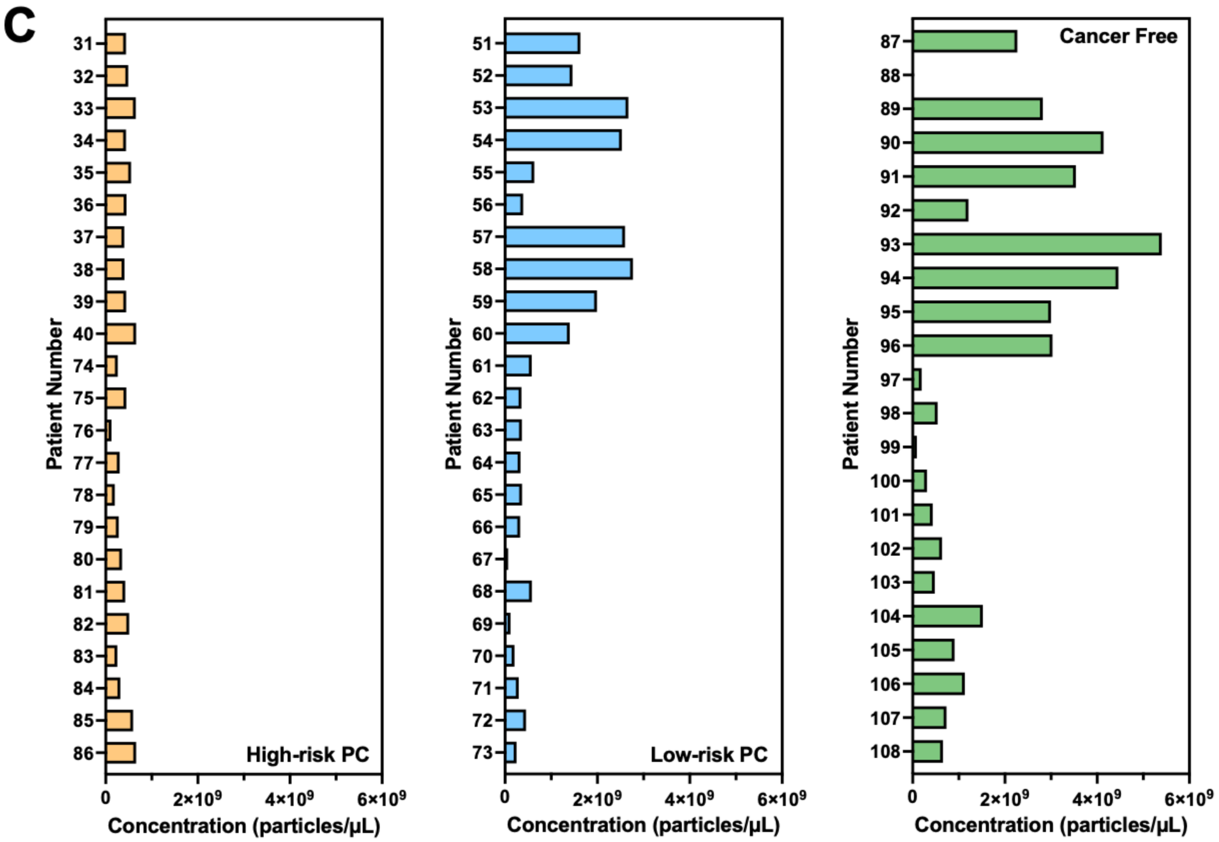


**Supplementary Figure S3. Analysis of EV-Trop2 ELISA data. (A)** Raw absorbance value ($OD_{450}$) from ELISA readout of each patient. **(B)** Bar graphs represent the normalized absorbance value ($OD_{450}$) from ELISA readout of each patient, with blank values subtracted. Line graph represents the corresponding EV-Trop2 levels (pg) based on the calibration curves in Supplementary Figure S1D. Error bar is standard deviation. **(C)** EV concentration of each patient (particles/µL) measured using NTA. Final EV-Trop2 levels (Figure 2C) were normalized to the number of EVs used for the ELISA, which was calculated by the product of EV concentration and volume used (5 µL) for all the samples. Detailed calculation can be found in the Methods section.

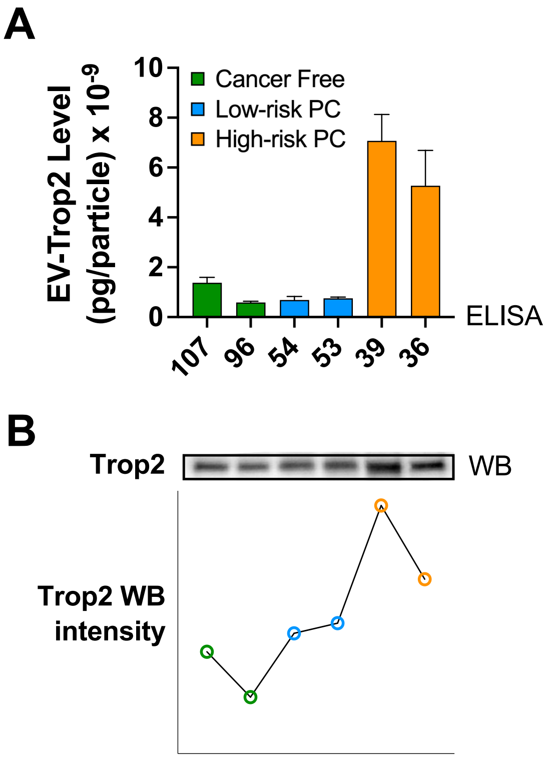


**Supplementary Figure S4. Comparative analysis of serum-derived EV-Trop2 levels employing ELISA and Western Blot (WB).** Selected patients from each group had their EV-Trop2 levels assessed using **(A)** ELISA and **(B)** Western Blot. Error bars are standard deviations.

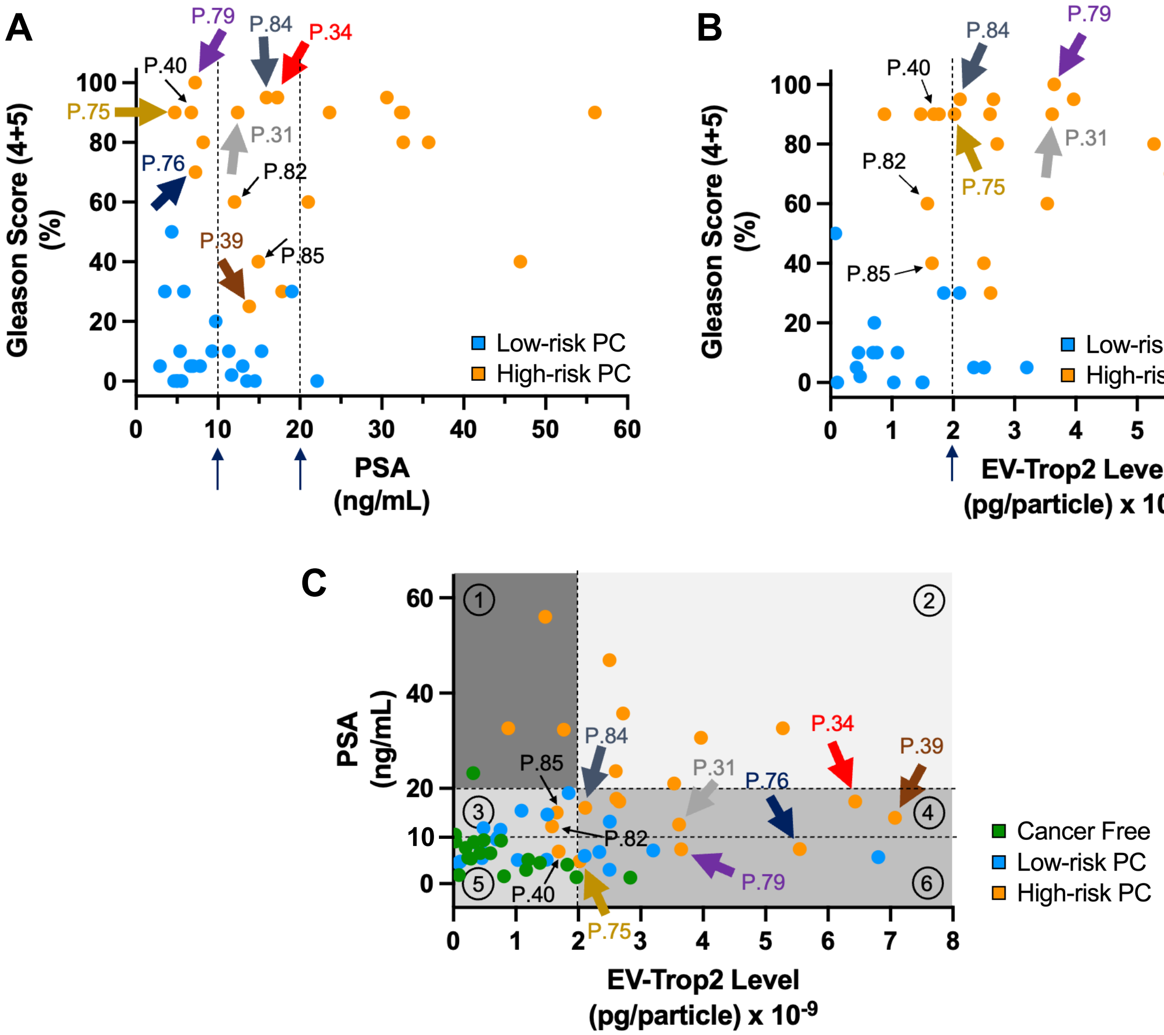


**Supplementary Figure S5. Illustration of EV-Trop2 levels using clinical risk guidelines. (A)** AUA and PCF guidelines use PSA levels to classify prostate cancer risk group as low-risk PC (<10 ng/mL), intermediate-risk (10–<20 ng/mL), and high-risk (≥ 20 ng/mL). Their PSA levels are correlated with %Gleason Score (Grade 4+5). **(B)** Correlation between %Gleason Score (Grade 4+5) and EV-Trop2 levels. **(C)** Correlation between PSA and EV-Trop2 levels. Large arrows indicate instances of biopsy-confirmed high-risk PC patients with low PSA levels but elevated EV-Trop2 levels, highlighting the potential of combined analysis for improved risk stratification before considering tissue biopsy. Small arrows represent cases of high-risk PC patients that were not identified by either the PSA or EV-Trop2 test.

The current standards for risk assessment in prostate cancer includes prostate-specific antigen (PSA), digital rectal examination (DRE), and tissue biopsy. While PSA screening has been widely used, concerns have arisen regarding its lack of specificity to prostate cancer[1] and the fluctuation in its levels,[2,3] potentially leading to overdiagnosis and overtreatment.[4–8] The Prostate, Lung, Colorectal, and Ovarian (PLCO) cancer screening trials in the US, with follow-up periods of 7-10 years[9] and 10-13 years,[10] showed no evidence of a reduction in prostate cancer mortality from screening using PSA testing and DRE.

**A**

**PSA**

| | AUC | F1 Score | Precision | Recall |
|---|---|---|---|---|
| **Neural * Network** | 0.685 | 0.487 | 0.492 | 0.485 |
| Logistic Regression | 0.654 | 0.478 | 0.473 | 0.485 |
| Naive Bayes | 0.556 | 0.465 | 0.481 | 0.455 |
| SVM | 0.603 | 0.453 | 0.453 | 0.470 |
| AdaBoost | 0.591 | 0.449 | 0.445 | 0.455 |
| Gradient Boosting | 0.671 | 0.449 | 0.445 | 0.455 |
| kNN | 0.667 | 0.420 | 0.412 | 0.439 |
| SGD | 0.654 | 0.420 | 0.350 | 0.530 |
| Random Forest | 0.623 | 0.378 | 0.382 | 0.379 |

Risk Group Stratification
Classifier Metrics (Neural Network)

| (%) | High-risk PC | Low-risk PC | Cancer Free |
|---|---|---|---|
| Sensitivity | 63.6 | 30.4 | 52.4 |
| Specificity | 86.4 | 62.8 | 73.3 |
| Accuracy | 78.8 | 51.5 | 66.7 |
| PPV | 70.0 | 30.4 | 47.8 |
| NPV | 82.6 | 62.8 | 76.7 |

**B**

**EV-Trop2 + PSA**

| | AUC | F1 Score | Precision | Recall |
|---|---|---|---|---|
| **Neural * Network** | 0.752 | 0.607 | 0.611 | 0.606 |
| SVM | 0.702 | 0.606 | 0.652 | 0.636 |
| kNN | 0.744 | 0.600 | 0.598 | 0.606 |
| Random Forest | 0.727 | 0.588 | 0.587 | 0.591 |
| Logistic Regression | 0.698 | 0.573 | 0.569 | 0.591 |
| Gradient Boosting | 0.736 | 0.531 | 0.533 | 0.530 |
| SGD | 0.700 | 0.471 | 0.398 | 0.591 |
| AdaBoost | 0.591 | 0.455 | 0.455 | 0.455 |
| Naive Bayes | 0.688 | 0.386 | 0.344 | 0.439 |

Risk Group Stratification
Classifier Metrics (Neural Network)

| (%) | High-risk PC | Low-risk PC | Cancer Free |
|---|---|---|---|
| Sensitivity | 72.7 | 43.5 | 66.7 |
| Specificity | 90.9 | 72.1 | 77.8 |
| Accuracy | 84.8 | 62.1 | 74.2 |
| PPV | 80.0 | 45.5 | 58.3 |
| NPV | 87.0 | 70.5 | 83.3 |

**Supplementary Figure S6. ROC curves assessing the performance of risk group classification using nine machine learning algorithms with EV-Trop2 and PSA data as input parameters.** Performance metrics comparing sample groups using **(A)** PSA alone versus **(B)** the combination of EV-Trop2 and PSA. Neural Network emerged as the top-performing model overall with the highest F1 score; thus it was used to generate the classifier metrics.

**Neural Network** is a class of models that utilize interconnected nodes (neurons) organized in layers to learn complex patterns from data.[11] They are particularly effective with high-dimensional data, such as images and audio, and can capture non-linear relationships.

In contrast, **Logistic Regression** models the probability of a binary outcome based on one or more predictor variables.[12] It is well-suited for datasets with continuous and categorical features but assumes a linear relationship between the independent variables and the log-odds of the dependent variable, which can limit its performance on more complex datasets.

**Naïve Bayes** classifiers are probabilistic models that apply Bayes' theorem under the assumption of feature independence.[13] They are particularly effective for text classification tasks, such as spam detection, where the features (words) are often independent of each other. While they are computationally efficient and work well with smaller datasets, their independence assumption may not hold true in practice, leading to suboptimal performance when features are correlated.

**Support Vector Machines (SVM)** are powerful supervised learning models that find the optimal hyperplane for classification tasks.[14] They can handle both linear and non-linear data through the use of kernel functions, making them versatile. SVMs excel in high-dimensional spaces, but they can be sensitive to the choice of kernel and require careful tuning of hyperparameters.

**AdaBoost** is an ensemble method that combines multiple weak classifiers to create a strong predictive model.[15] It works by iteratively adjusting the weights of incorrectly classified instances, focusing on hard-to-classify data. This method is effective with various base classifiers and performs well with both binary and multiclass classification problems. However, it can be sensitive to noisy data and outliers.

**Gradient Boosting** builds an ensemble of decision trees sequentially, optimizing for the loss function at each step.[16] It is highly flexible and can be adapted to different types of data, including continuous and categorical variables. While gradient boosting can achieve excellent performance, it may require more tuning and is prone to overfitting if not managed correctly.

**Random Forest** is another ensemble method that constructs multiple decision trees and merges their predictions for improved accuracy.[17] It handles both classification and regression tasks well and can work with mixed data types (continuous and categorical). Random forests are robust to overfitting, especially with larger datasets, but they can be less interpretable than simpler models like logistic regression.

The **k-Nearest Neighbors (kNN)** algorithm classifies data points based on the majority class among the nearest neighbors in the feature space.[18] It is a non-parametric method that works well with numerical and categorical data. However, kNN can become computationally expensive with large datasets and can be sensitive to the choice of distance metric and the value of k.

**Stochastic Gradient Descent (SGD)** is an optimization technique used to minimize a loss function iteratively.[19] It is particularly effective for training large-scale machine learning models and can handle different types of data. However, the performance of SGD depends on the choice of learning rate and can converge to suboptimal solutions if not properly tuned.

In summary, while these modeling approaches can be applied to various types of data, their effectiveness varies based on the data characteristics and the underlying assumptions of each model. Neural Network and Gradient Boosting excel in capturing complex patterns but require substantial data and tuning, whereas simpler models like Logistic Regression and Naive Bayes are more interpretable and efficient with smaller datasets. Ensemble methods like Random Forest and AdaBoost leverage the strengths of multiple models, offering robustness and flexibility across different tasks.

In the current study, we compared these approaches and preliminarily chose Neural Networks as an initial model since it ranked better than the other methods for risk classification. We do not claim that this model is the ultimate method to consider as we are aware that Neural Network ideally requires large data sets and prone to overfitting with sparse data. As such, we plan to carry out large cohort testing with adequately powered data sets in our future studies.

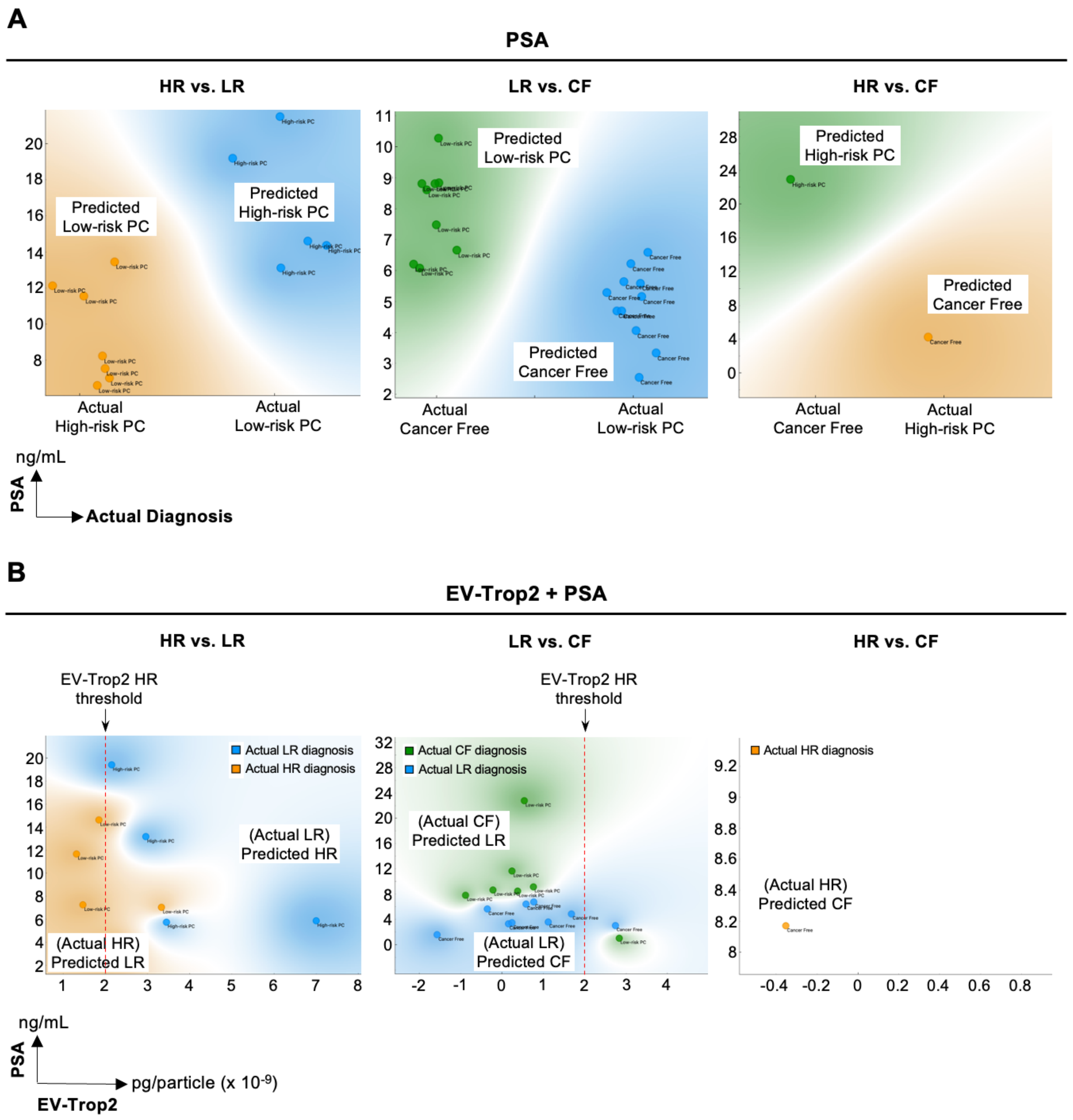


**Supplementary Figure S7. Error analysis to identify the features contributing to misclassification in our risk stratification models. (A)** PSA only model. **(B)** EV-Trop2 + PSA model. HR = high-risk PC, LR = low-risk PC, CF = cancer free

**PSA only** model misclassification analysis:

**HR vs. LR:**

- Misclassifications of HR primarily occur when PSA levels are below 14 ng/mL. In these cases, HR samples (orange dots) are misclassified as LR, despite their true status as HR. Conversely, some LR samples (blue dots) are misclassified as HR when their PSA levels exceed 14 ng/mL.
- As a reference, clinical guidelines typically classify HR cases as having PSA levels greater than 20 ng/mL. The misclassifications observed here suggest that using PSA alone for HR classification is suboptimal when PSA is below 14 ng/mL, and for LR classification when PSA exceeds 20 ng/mL.

**LR vs. CF:**

- CF samples (green dots) are misclassified as LR when PSA levels exceed 6 ng/mL, while LR samples are misclassified as CF when PSA falls below 7 ng/mL.
- According to clinical guidelines, CF is typically associated with PSA levels below 4 ng/mL, explaining why misclassifications are frequent in this borderline range.

**HR vs. CF:**

- Misclassification of HR as CF is observed when PSA levels are around 4 ng/mL—significantly lower than the HR threshold of 20 ng/mL.
- Similarly, CF samples are misclassified as HR when PSA levels are greater than 20 ng/mL, indicating that PSA alone may not effectively differentiate between HR and CF in certain cases.

**EV-Trop2+PSA** model misclassification analysis**:**

**HR vs. LR:**

- Misclassification of HR as LR occurs when EV-Trop2 levels fall below 2 pg/particle (x $10^{-9}$), a threshold set for HR identification (shown in Figure 2D and Figure 3). Conversely, LR samples are misclassified as HR when EV-Trop2 levels exceed 2 pg/particle (x $10^{-9}$).
- Interestingly, PSA levels (widely ranging from 4 to 20 ng/mL) do not show a clear trend in this analysis, suggesting that EV-Trop2 may be a reliable candidate marker than PSA alone in this classification. The combined EV-Trop2 and PSA model reduced the overall misclassification rate compared to the PSA-only model (*c.f.,* Supplementary Figure S7A above).

**LR vs. CF:**

- Misclassifications occur when EV-Trop2 levels are below 2–3 pg/particle (x $10^{-9}$). CF samples are misclassified as LR when PSA levels are higher than 6 ng/mL and EV-Trop2 levels are below 3 pg/particle (x $10^{-9}$). Conversely, LR samples are misclassified as CF when PSA levels are lower than 7 ng/mL and EV-Trop2 levels remain below 3 pg/particle (x $10^{-9}$).

**HR vs. CF:**

- Misclassification of HR as CF happens when PSA levels are around 8 ng/mL (significantly lower than the HR guideline of 20 ng/mL) and EV-Trop2 levels are much lower than 2 pg/particle (x $10^{-9}$).

**A**

**PSA**

| | AUC | Accuracy | F1 Score | Prec | Recall |
|---|---|---|---|---|---|
| **Naïve * Bayes** | 0.708 | 0.733 | 0.733 | 0.733 | 0.733 |
| Neural Network | 0.767 | 0.733 | 0.731 | 0.740 | 0.733 |
| Logistic Regression | 0.771 | 0.711 | 0.711 | 0.711 | 0.711 |
| Random Forest | 0.700 | 0.667 | 0.666 | 0.667 | 0.667 |
| SVM | 0.713 | 0.667 | 0.665 | 0.669 | 0.667 |
| Stochastic Gradient Descent | 0.640 | 0.644 | 0.631 | 0.662 | 0.644 |
| AdaBoost | 0.600 | 0.600 | 0.600 | 0.600 | 0.600 |
| kNN | 0.702 | 0.600 | 0.600 | 0.600 | 0.600 |
| Gradient Boosting | 0.690 | 0.578 | 0.577 | 0.578 | 0.578 |

Recurrence Prediction Classifier Metrics (Naïve Bayes)

| | |
|---|---|
| Sensitivity | 72.7% |
| Specificity | 73.9% |
| Accuracy | 73.3% |
| PPV | 72.7% |
| NPV | 73.9% |

**B**

**EV-Trop2 + PSA**

| | AUC | Accuracy | F1 Score | Prec | Recall |
|---|---|---|---|---|---|
| SVM | 0.806 | 0.800 | 0.800 | 0.800 | 0.800 |
| Gradient Boosting | 0.828 | 0.800 | 0.799 | 0.806 | 0.800 |
| Stochastic Gradient Descent | 0.798 | 0.800 | 0.799 | 0.805 | 0.800 |
| **Naïve * Bayes** | 0.804 | 0.778 | 0.778 | 0.780 | 0.778 |
| Neural Network | 0.834 | 0.778 | 0.777 | 0.780 | 0.778 |
| Logistic Regression | 0.824 | 0.756 | 0.755 | 0.756 | 0.756 |
| Random Forest | 0.778 | 0.733 | 0.733 | 0.736 | 0.733 |
| AdaBoost | 0.711 | 0.711 | 0.711 | 0.712 | 0.711 |
| kNN | 0.736 | 0.711 | 0.709 | 0.714 | 0.711 |

Recurrence Prediction Classifier Metrics (Naïve Bayes)

| | |
|---|---|
| Sensitivity | 81.8% |
| Specificity | 73.9% |
| Accuracy | 77.8% |
| PPV | 75.0% |
| NPV | 81.0% |

**Supplementary Figure S8. ROC curves assessing the performance of recurrence prediction using nine machine learning algorithms with EV-Trop2 and PSA data as input parameters.** Performance metrics using **(A)** PSA alone versus **(B)** the combination of EV-Trop2 and PSA. Naïve Bayes was chosen as one of the top-performing models overall to generate the classifier metrics.

**A**

| Risk Group | ID# | Pre-op PSA (ng/mL) | GR (4+5) Gleason Score (%) | Status |
|---|---|---|---|---|
| Cancer Free | 87 | 9.11 | N.A. | Healthy |
| n = 21 | 89 | 8.724 | N.A. | Healthy |
| | 90 | 6.4 | N.A. | Healthy |
| | 91 | 5.23 | N.A. | Healthy |
| | 92 | 10.27 | N.A. | Healthy |
| | 93 | 23.18 | N.A. | Healthy |
| | 94 | 8.83 | N.A. | Healthy |
| | 95 | 7.49 | N.A. | Healthy |
| | 96 | 6.36 | N.A. | Healthy |
| | 97 | 2.87 | N.A. | Healthy |
| | 98 | 3.96 | N.A. | Healthy |
| | 99 | 5 | N.A. | Healthy |
| | 100 | 2.57 | N.A. | Healthy |
| | 101 | 1.33 | N.A. | Healthy |
| | 102 | 1.53 | N.A. | Healthy |
| | 103 | 1.78 | N.A. | Healthy |
| | 104 | 5.42 | N.A. | Healthy |
| | 105 | 6.79 | N.A. | Healthy |
| | 106 | 9.01 | N.A. | Healthy |
| | 107 | 4.34 | N.A. | Healthy |
| | 108 | 1.24 | N.A. | Healthy |

**B**

| Risk Group | ID# | Pre-op PSA (ng/mL) | GR (4+5) Gleason Score (%) | Status |
|---|---|---|---|---|
| Low Risk PC | 51 | 15.3 | 10 | Cure |
| n = 23 | 52 | 9.7 | 20 | Cure |
| | 53 | 11.3 | 10 | Cure |
| | 54 | 9.26 | 10 | Cure |
| | 55 | 6.63 | 5 | Cure |
| | 56 | 2.9 | 5 | Cure |
| | 57 | 7.8 | 5 | Cure |
| | 58 | 11.64 | 2 | Cure |
| | 59 | 4.62 | 0 | Cure |
| | 60 | 4.33 | 50 | Cure |
| | 61 | 14.49 | 0 | Cure |
| | 62 | 5.01 | 0 | Cure |
| | 63 | 4.95 | 0 | Cure |
| | 64 | 13.51 | 0 | Cure |
| | 65 | 5.41 | 0 | Cure |
| | 66 | 19 | 30 | Cure |
| | 67 | 3.49 | 30 | Cure |
| | 68 | 5.35 | 10 | Cure |
| | 69 | 5.54 | 0 | Cure |
| | 70 | 13 | 5 | Cure |
| | 71 | 22.08 | 0 | Cure |
| | 72 | 5.81 | 30 | Cure |
| | 73 | 6.95 | 5 | Cure |

**C**

| Risk Group | ID# | Pre-op PSA (ng/mL) | GR (4+5) Gleason Score (%) | Status |
|---|---|---|---|---|
| High risk PC n = 22 | 31 | 12.38 | 90 | Fail |
| | 33 | 32.3 | 90 | Fail |
| | 34 | 17.22 | 95 | Fail |
| | 35 | 21 | 60 | Fail |
| | 36 | 32.6 | 80 | Fail |
| | 37 | 46.9 | 40 | Fail |
| | 38 | 30.6 | 95 | Fail |
| | 39 | 13.8 | 25 | Fail |
| | 40 | 6.72 | 90 | Fail |
| | 74 | 35.7 | 80 | Fail |
| | 75 | 4.7 | 90 | Fail |
| | 76 | 7.23 | 70 | Fail |
| | 77 | 8.17 | 80 | Fail |
| | 78 | 17.22 | 95 | Fail |
| | 79 | 7.19 | 100 | Fail |
| | 80 | 23.6 | 90 | Fail |
| | 81 | 56 | 90 | Fail |
| | 82 | 12 | 60 | Fail |
| | 83 | 17.8 | 30 | Fail |
| | 84 | 15.9 | 95 | Fail |
| | 85 | 14.9 | 40 | Fail |
| | 86 | 32.6 | 90 | Fail |

**Supplementary Table S1. Patient information categorized by risk groups**: **(A)** cancer free (n = 21), **(B)** low-risk PC (n = 23), and **(C)** high-risk PC (n = 22). Data includes pre-operative PSA level (ng/mL), %Gleason Score (Grade 4+5), and clinical status; classified as healthy (biopsy-confirmed absence of prostate cancer), cure (no biochemical recurrence at least 5 years after radical prostatectomy), and fail (recurrence after radical prostatectomy).
